# Deriving Mendelian Randomization-based Causal Networks of Brain Imaging Phenotypes and Bipolar Disorder

**DOI:** 10.1101/2024.12.12.24318953

**Authors:** Shane O’Connell, Brielin C. Brown, Dara M. Cannon, Pilib Ó Broin, David A. Knowles, Nadine Parker, Dag Alnæs, Lars T. Westlye, Saikat Banerjee, Leila Nabulsi, Emma Corley, Ole A. Andreassen, Niamh Mullins

## Abstract

Neuroanatomical variation in individuals with bipolar disorder (BD) has been previously described in observational studies. However, the causal dynamics of these relationships remain unexplored. We performed Mendelian Randomization of 297 structural and functional neuroimaging phenotypes from the UK BioBank and BD using genome-wide association study summary statistics. We found 28 significant causal relationship pairs after multiple testing corrections containing BD as a term, 27 of which described neuroimaging phenotype effects on BD. We applied an inverse sparse regression algorithm to estimate the direct effect of phenotypes conditional on all other causal effects, finding that white matter tract phenotypes have larger absolute effects on BD than *vice versa*. We found that white matter phenotypes have significantly larger out-degrees than non-white matter tract phenotypes, and that the effect of neuroimaging variation on BD is larger than *vice versa*. Our results provide support for the hypothesis that neuroanatomical variation, specifically in white matter tracts such as the superior and inferior longitudinal fasciculi, is a cause rather than a consequence of BD.

## Introduction

Bipolar disorder (BD) is a heritable mood disorder with a population prevalence of ∼2% worldwide ^1^. Its presentation consists of recurrent (hypo)mania, depression, and often psychotic symptoms ^2^. Owing to its considerable public health burden and incidence in families, its etiology has become the focus of intense research ^3,4,5^. Specifically, genome-wide association studies (GWAS) of BD have helped to characterize a fraction of its common genetic architecture, with over 60 genome-wide significant (GWS) loci identified in the largest published study to date ^6^.

There has also been a focus on describing neuroanatomical and neurofunctional variation in BD ^7,8^. Phenotypes indexing general brain variation are heritable according to previous GWAS of large populations ^9^. Observational magnetic resonance imaging (MRI) studies have described volumetric differences between individuals with BD and controls in regions such as the prefrontal cortex ^10–12^. Models of BD pathophysiology have posited that onset and disease progression likely emanate from changes in the structure and function of brain regions involved in emotional regulation ^10–12^. However, estimating directional causality is a challenging task with observational data.

Mendelian randomization (MR) methods use robustly associated genetic variants as instruments to estimate the causal relationship between two variables, eliminating certain types of confounding under specific assumptions ^13^. In practice, MR can be performed with GWAS summary statistics using a two-sample framework ^14^. The proliferation of GWAS using large-scale epidemiological resources such as the UK Biobank (UKB) has facilitated a range of causal analyses using MR methods across multiple phenotypes, including psychiatric conditions ^14–17^. Recent neuroimaging releases have resulted in systematic GWAS of MRI-based phenotypes indexing neuroanatomical and neurofunctional variation ^9,18^. These publicly available summary statistics have enabled two-sample MR studies to investigate the relationship between brain region variation and psychiatric conditions ^19^. However, previous work has focused on investigating specific brain phenotype categories ^20,21^, or has conditioned phenotype selection on significant genetic correlation between brain regions and psychiatric phenotypes (determined by the Linkage-disequilibrium Score Regression method ^19,22^). This may remove important phenotypes, as MR estimates are not dependent on genome-wide genetic correlation between two traits. This is because MR methods make use of independent variants robustly associated with the exposure, and the relationship between these effect estimates in the exposure and outcome may not always be correlated with the strength of the genome-wide genetic correlation between the exposure and outcome. Thus, a systematic analysis of the causal relationship between neuroimaging variables and BD remains unexplored. Additionally, the causal relationship between brain regions has not been previously described, which could be of interest in the context of psychiatric conditions. Furthermore, the utility of causal estimates in a predictive capacity for BD has not been tested.

Here, we carry out MR experiments to estimate the causal relationship between brain imaging variables from the UKB and BD. Additionally, we apply a novel causal network estimation method, inverse sparse regression, to account for the covariance structure between multiple correlated causal effects, yielding an estimate of the direct causal effects linking brain region phenotypes and BD in both directions ^23^. Using these causal estimates, we derive a risk score for BD using neuroimaging data in two independent cohorts and assess its predictive ability.

## Methods

### GWAS summary statistics and phenotype selection

Figure 1 displays the study workflow. We downloaded GWAS summary statistics of 3,929 brain imaging phenotypes in the UK Biobank^18^. These GWAS were carried out per-phenotype in unrelated individuals of European descent in discovery (n=∼22k) and replication (n=∼11k) sets separately^18^. We obtained PGC BD summary statistics comprising 41,917 BD cases and 371,549 controls of European ancestries^6^. We used Generalized Summary Mendelian Randomization (GSMR2) as our primary MR analysis method and as recommended, we considered phenotypes with at least 10 quasi-independent instruments for analysis ^24,25^. To do so, we identified phenotypes with at least 10 GWS SNPs via linkage disequilibrium (LD)-based clumping on the discovery GWAS summary statistics using the following parameters in PLINK 1.9^26^: LD *r*^2^ *>* 0.001, window 10,000 kb, *P* ≤ 5*e*−8. We used a subset of 16,886 individuals of European descent from the Haplotype Reference Consortium as an LD reference panel^26,27^. We did not consider phenotypes describing imaging quality control.

**Figure 1.**
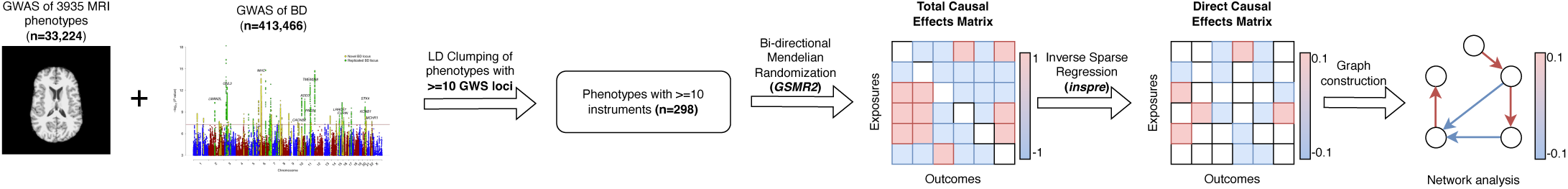
Study workflow. Figure made using draw.io. Summary statistics were assembled from the latest BD GWAS ^6^ and GWAS of over 3000 brain phenotypes ^18^. Clumping was performed using PLINK1.9 ^26^. We utilized GSMR2 as our primary causal estimation method using instruments from the discovery samples of the UKB phenotypes as exposures ^24^. inspre network analysis was performed using Cytoscape and the networkx package.

Our phenotype selection strategy was not conditioned on genetic correlation (*rg*), as MR causal estimates and *rg* estimates are not always correlated ^23,28^. Therefore, removing phenotypes with significant genetic correlations with other phenotypes may lead to information loss. We calculated pairwise *rg* estimates between all phenotypes (298 choose 2 = 44,253 tests) using LD-score regression ^22,29^, applying a Benjanini-Hochberg correction for multiple testing, and constructed a genetic correlation matrix for comparison with our causal effect matrix (Fig. S1).

### *GSMR2* analysis

All MR analyses were carried out in compliance with the STROBE-MR guidelines ^30^. We performed 298 choose 2 sets of forward and reverse MR tests (88,506 tests) using GSMR2 with the following parameters: heterogeneity in dependent instruments (HEIDI) p-value threshold = 0.01, LD *r*^2^ threshold = 0.05, LD FDR threshold = 0.05, including 297 brain imaging phenotypes and BD. For estimating the causal effect of brain phenotypes on each other, we used SNPs from the discovery sample for exposures and SNPs from the replication sample for outcomes, to avoid potential bias arising from sample overlap ^31^. The HEIDI test detects SNPs which are pleiotropic between the exposure and outcome, thus violating the instrumental assumptions of MR, and these were removed prior to causal estimation. We applied a Benjamini Hochberg correction to the resultant p-value matrix and considered MR tests with BD as a term at *Q <* 0.05 and *Q <* 0.01 for further investigation (where *Q* is the FDR-corrected p-value). If exposure instruments were not present in sufficient quantities in outcome summary statistics (<10), causal effects were not estimated. A 298×298 matrix of exposures (rows) by outcomes (columns) was populated using the *β* coefficients of all MR tests, representing our total causal effects (TCE) matrix. We tested for a difference in means in the absolute effect of BD on every other phenotype in the matrix versus the absolute effect of every phenotype in the matrix on BD using an independent t-test. Brain imaging phenotypes were split into 13 categories as described in the original publications and detailed further in the results section ^18,31^. We further tested for a difference in means in the absolute effect on BD per phenotypic category versus the absolute effect of BD on that phenotype category using an independent t-test, with a Bonferroni correction for the 13 categories tested (P=0.05/13=3.84e-3). We measured the correlation between every phenotype’s genetic correlation profile and their causal effects as exposures. Pairs with FDR-significant (*Q <* 0.01) p-values containing BD were compared to FDR-significant (*Q <* 0.01) MR pairs containing BD as a term.

### Sensitivity Analyses

FDR-significant exposure-outcome pairs including BD as a term underwent several sensitivity analyses. These included a leave-one-instrument-out analysis, confounder-associated instrument removal, and multiple MR methods to assess robustness under different modeling assumptions. The panel of MR methods considered included the inverse variance weighted, simple mode, weighted mode, Egger regression, and weighted median methods ^32,33^. We used the *TwoSampleMR* package to apply these methods using SNPs identified as valid instruments by *GSMR2* ^15^. We plotted the resultant causal estimates and their confidence intervals to determine the consistency of causal effects (Figures S2-S15). To identify phenotype pairs where causal estimates across SNPs were heterogeneous, we performed leave-one-instrument-out analyses. This involved running an inverse variance weighted regression to obtain causal estimates while removing one valid instrument at a time ^34,35^. We repeated this operation per FDR-significant phenotype and plotted resultant *β* values, noting where test statistics lost statistical significance (Figure S16).

We identified a panel of nine phenotypes that were potential confounders of the exposure-outcome relationship defined in the main BD GWAS study for more focused instrument exclusion experiments ^6^. These included problematic alcohol use disorder, smoking initiation, cigarettes per day, drinks per week, morningness, insomnia, and educational attainment ^36–42^. We obtained summary statistics for each phenotype from *GWASCatalog*^43^. Exposure instruments that were also significant associations (*P* ≤ 5*e*−8) of a potential confounder were removed and *GSMR2* was rerun. We plotted the difference in *β* values and p-values before and after confounder-associated SNP removal to examine if the estimates were robust (Figure S17).

### *inspre* analysis

We carried out inverse sparse regression using the *inspre* package to estimate direct causal effects (DCE) from total causal effects ^23^. Briefly, this operation seeks to derive a precision matrix-like quantity representing the conditional dependencies between input entries, yielding a sparse output graph where the covariance structure between inputs has been accounted for. This collapses to a modified graphical lasso procedure. Further details on the algorithm can be found in the main text and supplemental note of the original paper by Brown and colleagues ^23^. We chose stable output solutions using the stability approach to regularization strength selection method ^44^. The stability metric, termed *D̂*, represents the average probability of edge inclusion in outputs under random re-samplings of the input across different penalty parameters (*λ*). Further details on the implementation of this method can be found in the primary methods paper ^23^. Stable solutions were classified as those with *D̂* values below 0.05 ^44^.

Multiple stable output solutions can exist with varying levels of numerical sparsity. We iterated over several stable solutions across a range of *λ* values using 10-fold cross validation to obtain a range of potentially valid output graphs. We plotted the correlation between candidate solutions satisfying stability criteria (*D̂* ≅ 0.05) (Figures S18-S20). In sparse solutions, we counted the number of times a phenotype was a non-zero effector of BD (Figure S20). Using our specified output solutions, we repeated statistical tests carried out in the TCE matrix, testing for a difference in means between the absolute effect of BD on all phenotypes and the absolute effect of all phenotypes on BD using an independent t-test. We tested for a difference in means in the same quantity per phenotypic category as previously described using 13 independent t-tests which were Bonferroni-corrected for multiple testing.

### Network construction

We created subnetworks from our candidate solutions using several filtering conditions. Firstly, we ranked the top 20 phenotypes with the largest absolute DCE effects on BD and created a 21 × 21 matrix (20 phenotypes plus BD) of DCE estimates. We created a directed network from this matrix using the *networkx* package, interpreting the input as a weighted adjacency matrix. We then removed all edges with absolute weights below one standard deviation of the global mean DCE effect and visualized our network in *Cytoscape*. We carried out this process for selected stable solutions. For the selected network solutions, we also visualized the same networks using TCE and *rg* estimates. We created a standalone web application summarizing all information for these edges available for download in the Supplementary Note.

For network visualization and experiments, we grouped phenotypes into white matter phenotypes (any with ‘white matter’ or ‘WM’ in their descriptions ^18,23^) and grey matter structural phenotypes. We tested for a difference in out-degree between the categories across selected stable solutions to determine which category had more direct influence on the wider network using an independent t-test. Out degree was calculated as the number of non-zero targets of a phenotype as an exposure in the network.

### BD prediction using causal estimates

We assessed the predictive capabilities of our causal estimates for BD in two separate cohorts of clinically defined BD participants and controls^12^. The first cohort had a total sample size of 100, with 44 BD cases and 56 controls; the second cohort had a total sample size of 565, with 127 cases and 438 controls. The use of these cohorts was approved by local institutional review boards and ethics committees, and all study participants provided written informed consent. Full cohort demographic descriptions can be found in the Supplemental Note (Table S2).

Using subcortical volumetric data and fractional anisotropy (FA) measures available from our clinical samples, we matched 14 variables to brain imaging phenotypes in our direct causal matrices. We adopted a polygenic-risk-score-like approach, whereby we summed the product of our scaled causal weights and normalized neuroimaging measures per patient to derive a causal score metric, which we refer to as the causal *β* score ^12,45^. This procedure can be represented by the following:

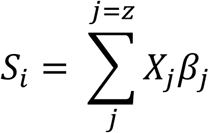

where *i* indexes the participant, *j* indexes the number of neuroimaging variables (*z*), *X* represents the neuroimaging measure, and *β* represents the causal estimate for that neuroimaging variable. This results in a causal *β* score *S* per individual. We fit the null model by performing a logistic regression of BD status against covariates, and the full model by regressing BD status against *S* plus covariates. We calculated the Nagalkerke’s pseudo-R2 variance explained for each model and subtracted that of the null model from the full model to obtain the variance explained by S, which was subsequently transformed to the liability scale to account for case ascertainment bias using a BD population prevalence of 2%^46,47^. We assessed model fit using an analysis of variance (ANOVA) model, comparing the null model to the full model. We determined an empirical p-value distribution for this quantity by randomly permuting BD and control status 1000 times and refitting null and full models, using an ANOVA to obtain a p-value for the fit ^48^. Significance was assessed by determining the proportion of tests with empirical p-values greater than the observed p-values. We carried out this procedure across cohorts using DCE weights to calculate *S* ^49^. We used the *r2redux* package to test for a significant difference between variance explained between *S* scores calculated using DCE and TCE weights ^50^. We performed all tests separately in each cohort and meta-analyzed the regression results using the dense DCE results with a random-effects model using the *metafor* R package ^51^.

## Results

### Causal relationships between neuroimaging measures and BD in TCE

We found 298 phenotypes with ≥ 10 GWS clumped instruments (297 brain imaging phenotypes plus BD). These phenotypes can be grouped into 13 phenotypic categories (Table S1) previously defined by Smith and colleagues ^18^. Briefly, these categories include regional and tissue volume (describing volumetric changes), WM tract ICVF (white matter tract intracellular volume fraction, estimated from the neurite orientation dispersion and density imaging model ^18,52^), cortical grey-white contrast, WM tract diffusivity (describing how freely water molecules can diffuse), cortical area, WM tract OD (white matter tract orientation dispersion, describing the orientation of diffusion), regional and tissue intensity, WM tract FA (white matter tract fractional anisotropy, describing the directionality of restricted diffusion), WM tract ISOVF (white matter tract isotropic or free water volume fraction), regional T2* (T2 intensity from susceptibility-weighted imaging, describing the amount of water content in a region), rsfMRI connectivity (resting state functional MRI connectivity, describing features from an independent components analysis (ICA) introduced by Smith and colleagues ^18^), WM tract MO (white matter tract diffusion tensor mode, describing whether or not multiple fibres are present in the high FA regions), and white matter hyperintensity volume. More details on phenotypic categories and the discussed constructs can be found in the Supplemental note of previous work performed by Elliott and colleagues ^9^. The best represented phenotypic group was regional and tissue volume phenotypes, with 71 phenotypes included in our analyses (Table S1). The median *SNP h*^2^ of included brain phenotypes was 0.3 (calculated using *ldsc* by Elliot and colleagues^18^). Cortical grey-white contrast phenotypes had the highest median heritability of any group category (median *SNP h*^2^ 0.33, Table S1).

Using our panel of 298 phenotypes, we carried out 88,506 forward/reverse MR experiments using GSMR2. After applying FDR correction, we found that 28,832 phenotype-phenotype pairs had significant causal estimates (Q<=0.01). The majority of significant causal pairs (28,805) were brain-phenotype on brain-phenotype causal estimates, with 27 brain phenotypes having an FDR-significant causal effect on BD (Figure 2A). Twenty of these significant exposures were white matter tract intracellular volume fraction (WM-ICVF) phenotypes. We found one significant causal pair featuring BD as an exposure at a relaxed threshold of Q<0.05 - the strength of the white-gray matter contrast in the right hemispheric insula (*β* = 0.094, 95% CI = 0.031 − 0.156, Q=0.011, Figure 2C, Table 1).

**Figure 2.**
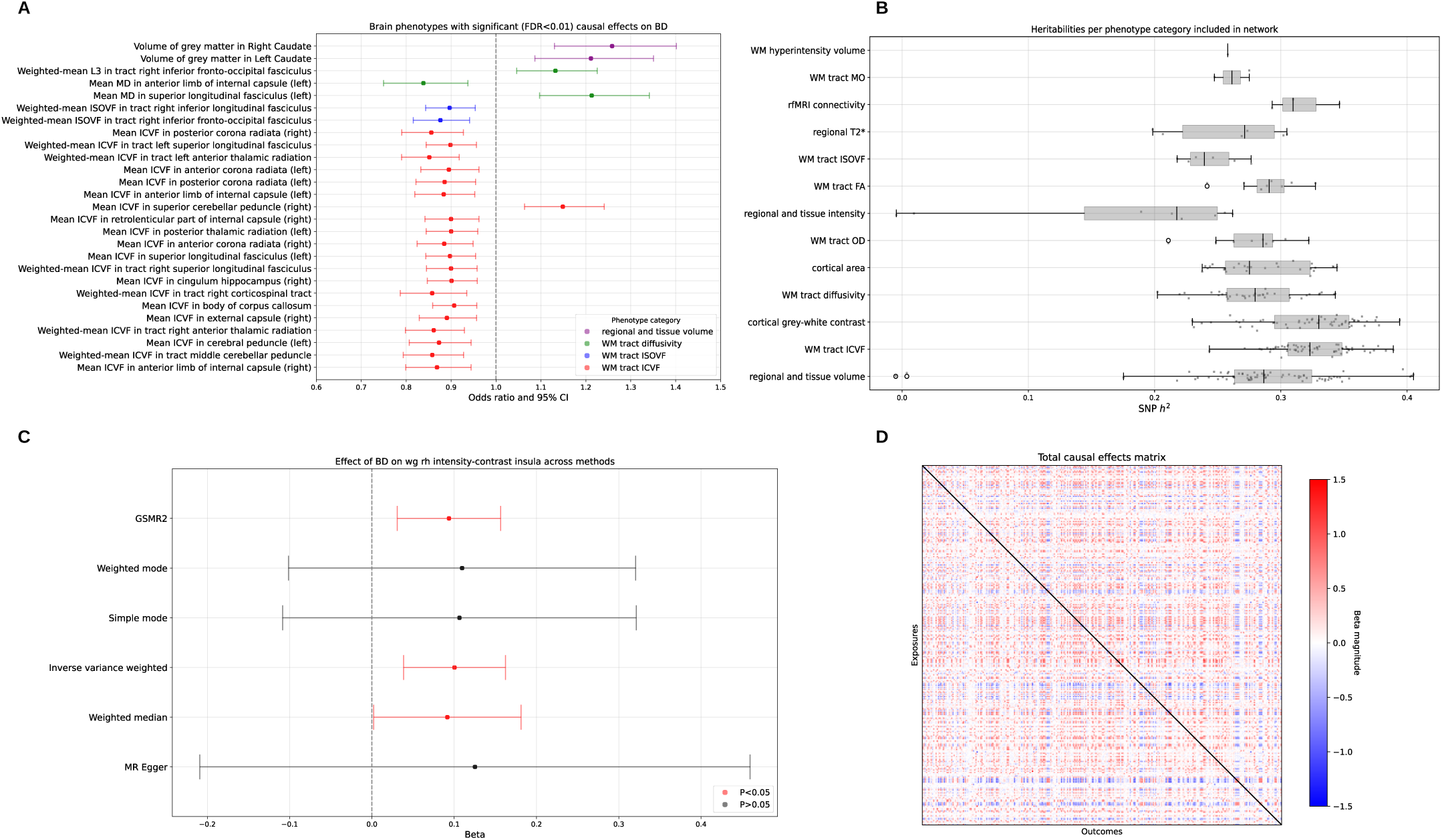
GSMR2 results and causal estimates. A) Plot of 27 causal relationships with FDR-significant test statistics at Q<0.01 containing BD as a term. Odds ratio and confidence intervals from GSMR2 are presented along the x-axis with phenotypes colored by their category. B) *SNPh*^2^estimates for phenotypes included in our MR analyses on the x-axis against phenotype category on the y-axis. Estimates were derived from the original paper which utilized ldsc to calculate heritability. From bottom to top, the y-axis is sorted by the number of phenotypes in that category in the network from largest to smallest. C) Plot of causal estimates and their associated confidence intervals for the effect of BD on the strength of the white-gray contrast in the right hemispheric insula across six MR methods. After FDR correction of GSMR2 p-values, this pair was found to have a Q value of 0.011. D) Matrix of total causal effects for all phenotypes included in our analyses. Exposures are presented along the rows, with the outcome of that exposure presented in the columns, making the matrix asymmetric.

**Table 1:**
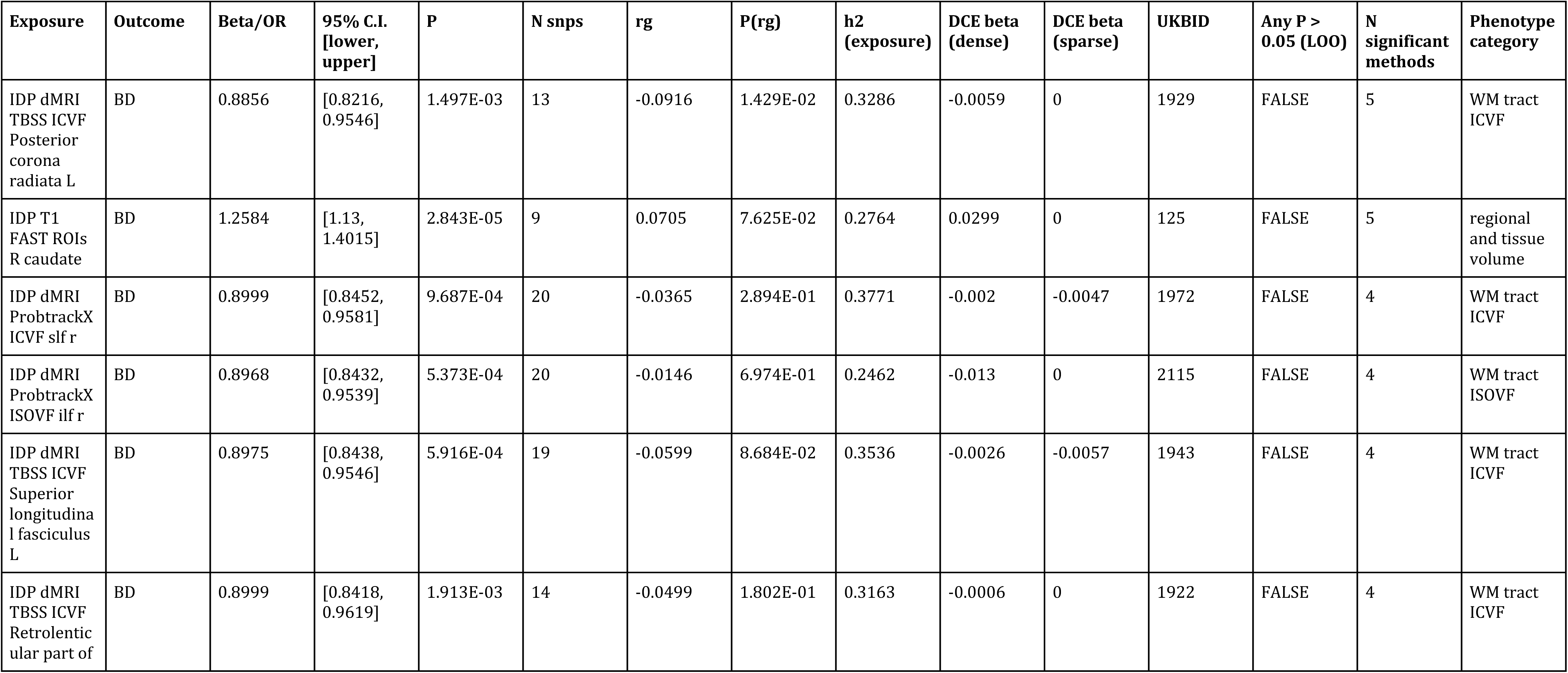
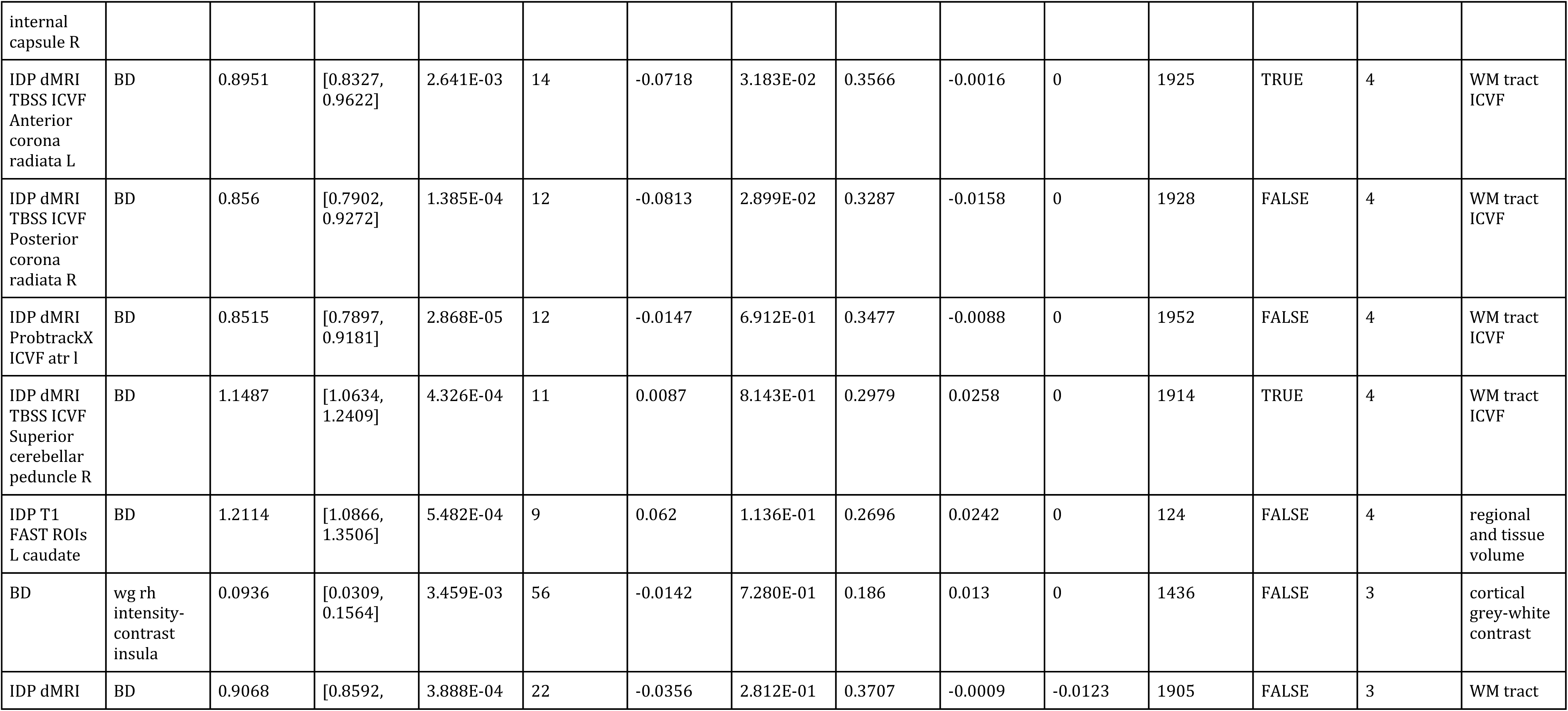
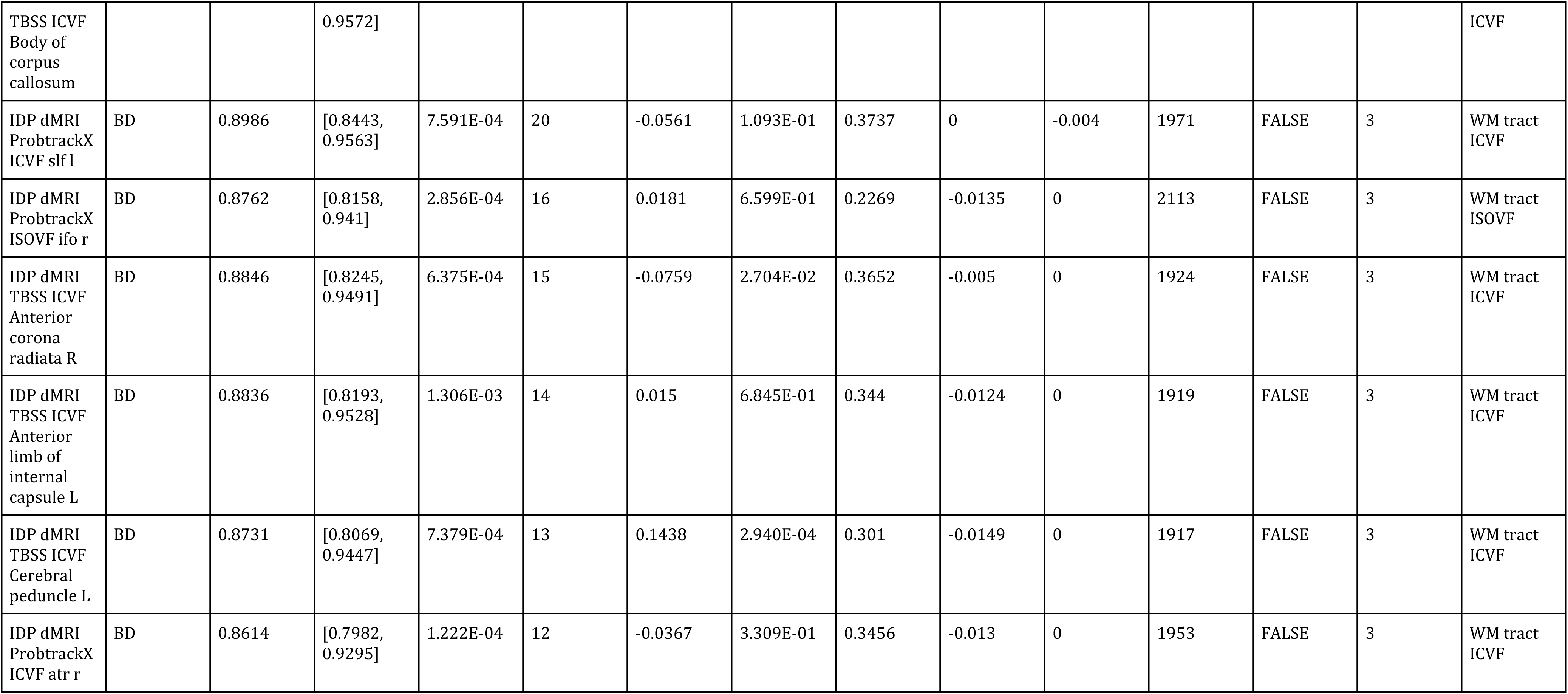
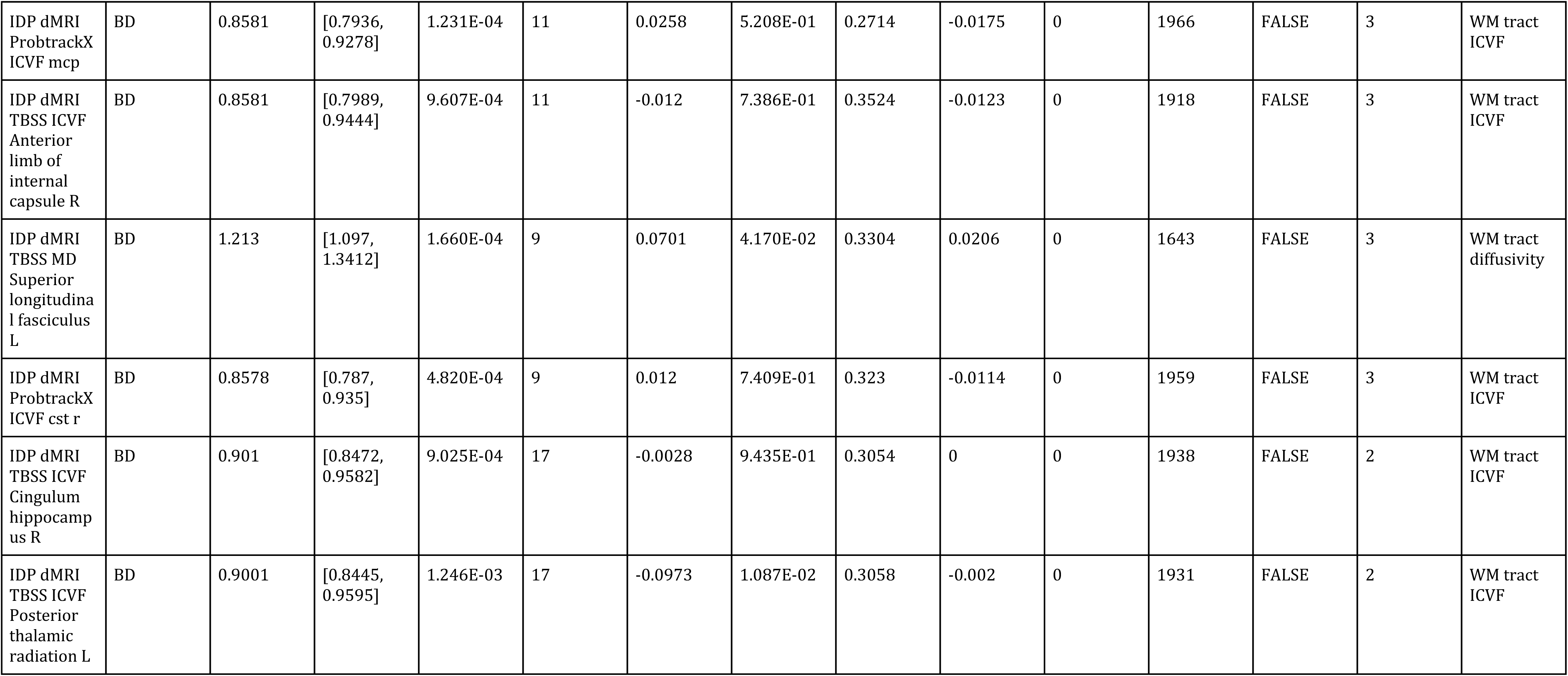
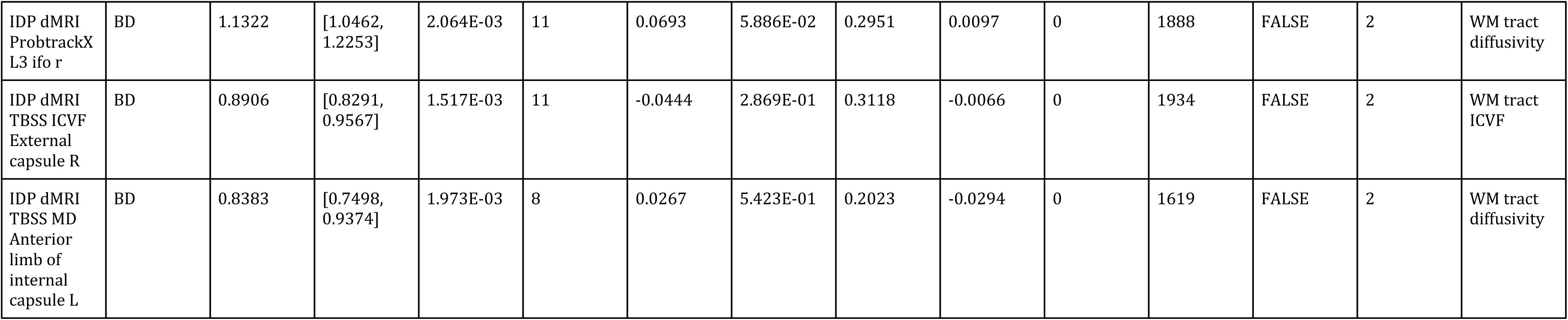
Information on 28 FDR-significant causal relationships between brain phenotypes and BD, including the UKBID, the results of sensitivity analyses, the phenotype category, the causal estimate from GSMR2 in either the odds ratio scale or *β* scale (depending on the data type of the outcome), the rg estimate, the DCE estimate (dense and sparse), the number of instruments, and the number of MR methods in which the pair was significant (out of 6 total methods). 95% C.I. stands for 95% confidence interval, with the lower and upper bounds presented accordingly. Any P>0.05 (LOO) denotes whether or not any test statistic lost significance after systematic leave-one-out SNP exclusion.

Our sensitivity analyses were focused on the 27 brain-phenotype-BD pairs found to be FDR-significant at Q<0.01 and the one BD-brain-phenotype pair found to be FDR-significant at Q<0.05. Our formal sensitivity analysis using *TwoSampleMR* resulted in 2 phenotypes with test statistics losing statistical significance after instrument removal (mean ICVF in the right superior cerebellar peduncle on BD; mean ICVF in the left anterior corona radiata on BD) (Figure S16). We found 9 SNPs overlapping between our panel of potential confounder GWAS significant hits and valid exposure instruments from our 28 pairs of interest. This resulted in 9 pairs requiring re-analysis with confounder-associated SNPs removed. All tests remained significant at *p* < 0.05 after SNP removal with minimal changes to estimated *β* values (Figure S17).

We found that 23/28 of our FDR-significant pairs had significant test statistics in at least three MR methods out of 6. For example, the standardized effect of BD on the strength of the white-gray matter contrast in the right hemispheric insula was found to be significant by *GSMR2,* inverse variance weighted regression, and the weighted median method. Two phenotypes were significant in at least 5 MR methods including *GSMR2*; mean ICVF in the left posterior corona radiata on BD, and the volume of the grey matter in the right caudate on BD. The effect of volume of the grey matter in the left caudate on BD had confidence intervals that did not contain zero across all methods, but p-values from the weighted and simple mode estimators were non-significant (Figure S2A). We found good correlation between *rg* estimates from *ldsc* and causal estimates per phenotype in the forward direction (median correlation of 0.84). After carrying out FDR correction, we found one phenotype with a significant (Q<0.01) genetic correlation test statistic with BD that was also identified as a significant causal exposure in our MR experiments: the mean ICVF in the left posterior corona radiata on BD. We constructed a TCE using the causal estimates from *GSMR2* (Figure 2D).

### Estimated direct causal associations between white matter phenotypes and BD

We found several candidate *inspre* solutions with instability metrics at our threshold of approximately 0.05. The correlation between 77 candidate solutions was high (median *ρ*=0.76, Figure S18). Most solutions were sparse (5 solutions with more than 20% non-zero entries, Figure S19). Amongst sparse solutions (72 solutions), four WM-ICVF phenotypes occurred as non-zero effectors of BD in at least 50% of outputs (mean ICVF in left superior longitudinal fasciculus, weighted-mean ICVF in right superior and left inferior longitudinal fasciculus, weighted-mean ICVF in forceps minor). Given the broad range of possible candidate solutions of varying numerical density, we focused on two stable output DCE matrices of differing non-zero percentages (Figure 3, Figure S19). Specifically, we chose a dense solution with 74% non-zero values and a sparse solution with 11% non-zero values (Figure S19). In our dense output, the largest effector of BD was the volume of the gray matter in the right caudate. In this matrix, we found that the mean causal effect of phenotypes on BD was statistically larger than *vice versa* (P=1.3e-8, Figure 4A). This result was mirrored in the TCE input, with phenotypes exerting a larger mean causal effect on BD than *vice versa* (P=2.76e-28). In our dense DCE solution, we found that in phenotypic categories with greater than 20 phenotypes, WM-ICVF phenotypes had a statistically larger estimated effect on BD than *vice versa* after Bonferroni correction (P=0.0012, Figure 4D). This result was also observed in the TCE input, with WM diffusivity, cortical grey-white contrast, and regional/tissue volume categories also found to be significant in the same direction (Figure 4C). We found that white matter phenotypes had a statistically larger out degree than non-white matter phenotypes in our dense solution (P=3.37e-12, Figure 4B). The correlation between our input TCE and dense DCE was 0.21 (Figure 3B). The top 20 causal effectors of BD in this solution contained 8 white matter phenotypes and 12 non-white matter phenotypes (Figure 5A).

**Figure 3.**
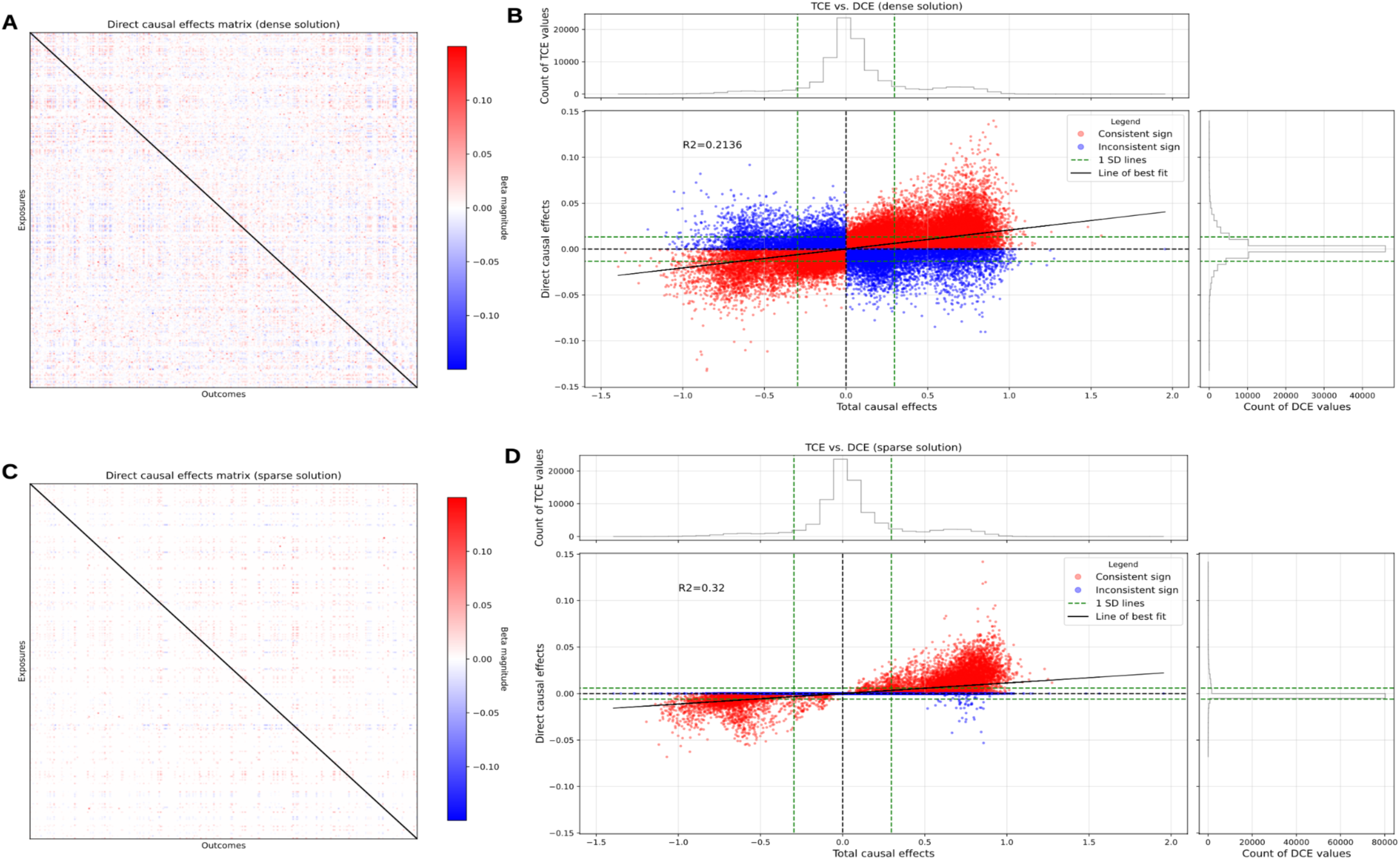
DCE estimates of input TCE. A) Matrix of direct causal effects from a stable graph solution estimated using inspre. Exposures are presented along the rows and outcomes along the columns, making the matrix asymmetric. This solution contains 74% non-zero values and a *D̂* estimate <=0.05. B) Correlation plot of the dense DCE solution from A against the input TCE from Figure 2D. Points are colored according to whether the sign of the causal estimate is consistent across input TCE and output DCE. Upper and right histograms describe the respective distributions of the TCE and dense DCE. Dotted green lines detail one standard deviation of the respective matrices. C) Matrix of direct causal effects from a stable graph solution estimated using inspre. Exposures are presented along the rows and outcomes along the columns, making the matrix asymmetric. This solution contains 11% non-zero values and a *D̂* estimate <=0.05. D) Correlation plot of the sparse DCE solution from C against the input TCE from Figure 2D. Points are colored according to whether the sign of the causal estimate is consistent across input TCE and output DCE. Upper and right histograms describe the respective distributions of the TCE and dense DCE. Dotted green lines detail one standard deviation of the respective matrices.

**Figure 4.**
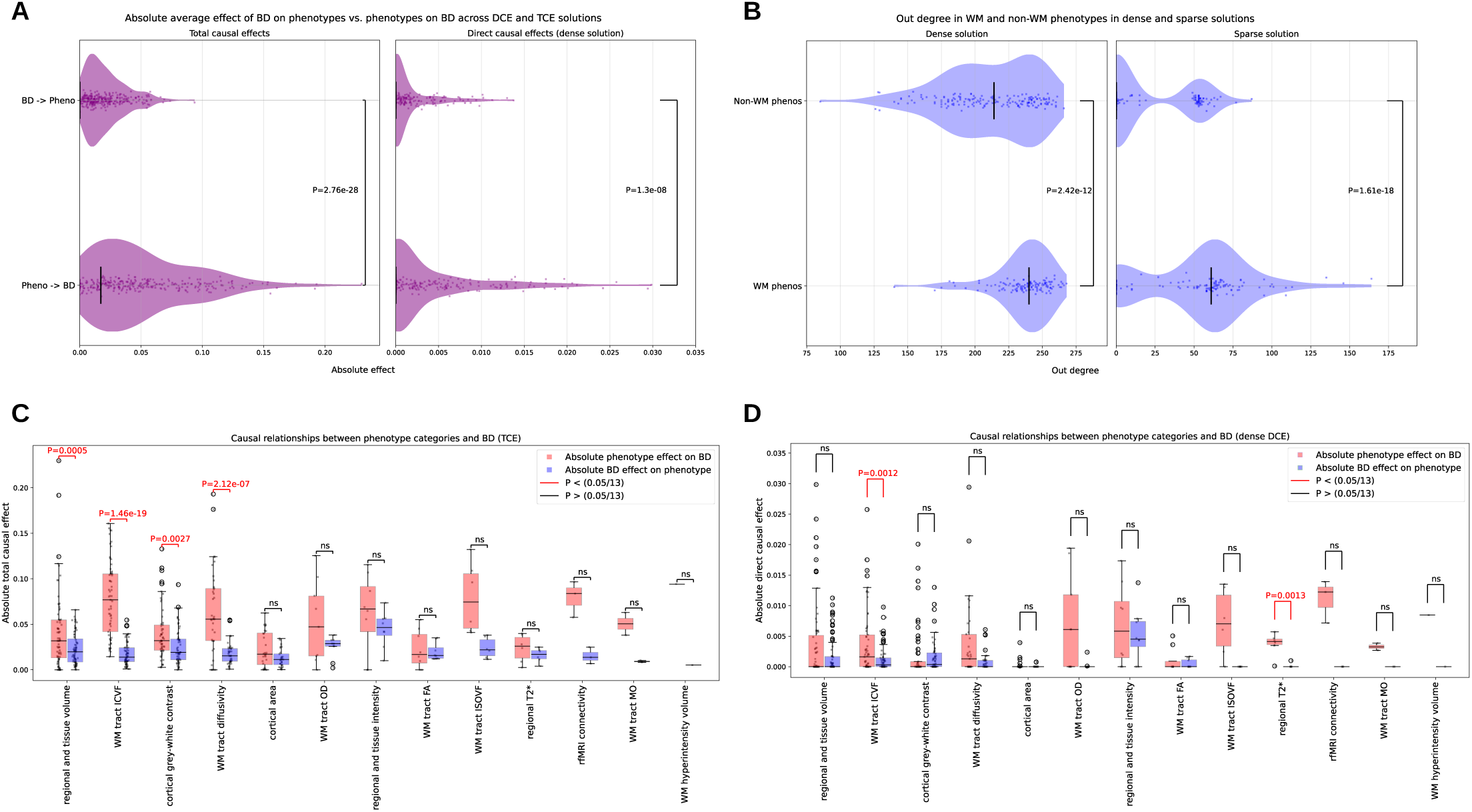
Causal effects on BD and neuroimaging phenotypes across DCE and TCE solutions. A) Violin plots of the absolute causal effect of BD on phenotypes and vice versa, where the absolute causal effect is detailed on the x-axis. Left panel uses the absolute causal effects from the TCE matrix and right panel uses the absolute causal effects from the dense DCE matrix. P-values were calculated from an independent t-test of means between both groups. B) Violin plots of out degree in different phenotypic categories across DCE solutions, where out degree is presented on the x-axis. Out degree is calculated as the number of non-zero connections from phenotypes of the specified category where the index phenotype acts as the exposure. P-values were calculated using an independent t-test of means. C) Absolute causal effect of BD on phenotypes and vice versa in the TCE stratified by phenotypic category. Independent t-tests were carried out within categories and p-values less than the Bonferroni-corrected significance level are colored red. D) Absolute causal effect of BD on phenotypes and vice versa in the DCE stratified by phenotypic category. Independent t-tests were carried out within categories and p-values less than the Bonferroni-corrected significance level are colored red.

**Figure 5.**
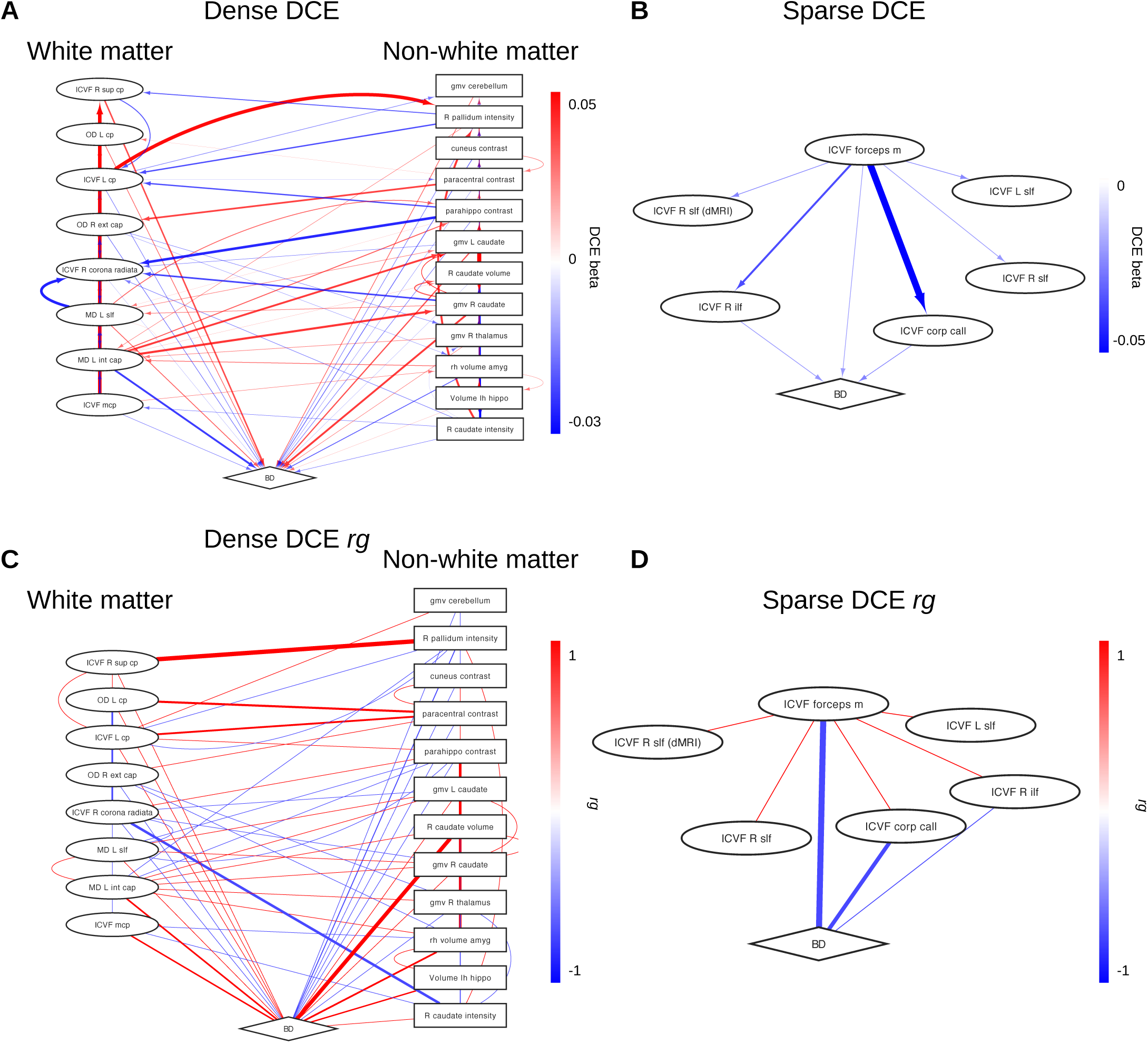
DCE estimates of input TCE. A) Network diagram of the top 20 exposures acting on BD in the dense DCE solution thresholded by one standard deviation. B) Network diagram of exposures acting on BD in the sparse DCE solution thresholded by one standard deviation. C) Network diagram of rg values between the interactions present in A. D) Network diagram of rg values between the interactions present in B.

Our sparse solution with 11% non-zero entries featured the mean ICVF in the corpus callosum as the largest effector of BD (Figure 5B). We found that all non-zero effectors of BD in this network were WM ICVF phenotypes. We found that white matter phenotypes had a statistically larger out degree than non-white matter phenotypes in this network (P=4.31e-19). Of the seven non-zero effectors of BD, five were phenotypes describing diffusivity in the superior or inferior longitudinal fasciculus, with the other two phenotypes describing diffusivity in the forceps minor and corpus callosum. The global mean causal estimate decreased from 0.065 to 0.001 in the dense DCE solution and decreased to 0.0008 in the sparse solution. We plotted the *rg* estimates for each of the aforementioned networks and found that the direction of effect was not always consistent (Figure 5C; 5D). Further, we constructed a standalone interactive *Cytoscape* web application summarizing Figure 5 in its entirety (Supplementary materials).

### Comparison of FDR-significant pairs across causal estimates and genetic correlation results

We observed that 3/28 FDR-significant causal pairs containing BD as a term had non-zero values across all considered experiments - ICVF in the corpus callosum, ICVF in the right hemispheric superior longitudinal fasciculus, and ICVF in the left hemispheric superior longitudinal fasciculus (Table 1). The direction of effect was consistent between all MR methods and both dense and sparse DCE solutions, with standardized unit increases in the phenotype values estimated to have a negative causal effect on BD. We also observed that the genetic correlation estimates were also negative, although the p-values of the test statistics were insignificant across all three phenotype pairs. Increased volume of the right caudate was estimated to have a significant causal effect on BD in 5 out of 6 considered MR methods (all models except Egger regression). Seven phenotype pairs had inconsistent signs with their *rg* estimates, including the effect of BD on the strength of the white-gray contrast in the right hemispheric insula. Seven out of 28 FDR-significant pairs had significant genetic correlation estimates, five of which described negative genetic correlations (Table 1). The range of *rg* values for the 7 significant pairs was −0.0973 to 0.1438, with the median *rg* found to be −0.0759.

### Prediction of BD status using causal estimates

We found that our causal *β* score calculated using DCE weights (direct causal *β* score) was significantly associated with age in our Galway cohort (P<0.05), and with age and sex in our Oslo cohort. Holding age constant, we estimated that a 1 s.d. increase in direct causal *β* score was associated with a 1.06 odds increase in BD risk (95% C.I. 0.69, 1.61; P=0.802) in the Galway population. The direct causal *β* score explained 0.0415% of variance (*R*^2^liability scale) in BD status and the area under the receiver operating characteristic curve (AUC) was 0.55. The empirical p-value of the direct causal *β* score was deemed insignificant after 1000 permutations of phenotype values across all individuals (empirical P=0.811).

In our Oslo testing population, we found that age and sex were significantly associated with our direct causal *β* score. Including both of these variables as covariates, we found that 1 s.d. unit increase in direct causal *β* score was associated with a 1.03 odds increase in BD status (95% C.I. 0.82, 1.29; P=0.805). In this cohort, the direct causal *β* score explained 0.01% of phenotypic variance on the liability scale. The empirical p-value of the direct causal *β* score was deemed insignificant after 1000 permutations of phenotype values across all individuals (empirical P=0.791). The AUC for this cohort was 0.60. After a random-effects meta-analysis, we found a nonsignificant association between direct causal *β* score and BD status across cohorts (OR = 1.03, 95% C.I. 0.85, 1.26; P=0.7359, Figure S21).

We found a non-significant difference between the variance explained using causal *β* scores calculated using either TCE or DCE weights (Galway two-sided P=0.856; Oslo two-sided P=0.644). We found that the liability *R*^2^ from causal *β* scores calculated using TCE weights was 0.009 in the Galway population, and 0.067 in the Oslo population.

## Discussion

We ran over 88 thousand MR analyses to examine the potential causal relationship between brain imaging phenotypes and BD, finding several significant causal pairs. Standardized unit increases in phenotypes indexing white matter microstructural variation typically had negative causal effects on BD status. Previous BD neuroimaging and endophenotype research has focused on observed white matter microstructural differences, with a large mega-analysis reporting significantly decreased FA in the body of the corpus callosum ^53,54^. Additionally, the mean ICVF in the corpus callosum was found to be one of three FDR-significant phenotypes with non-zero values in both selected DCE solutions. Whether or not decreased FA (used as a proxy for tract microstructural organization, with lower values implying less directionally restricted diffusion) and decreased ICVF (representing less axonal tissue, or decreased neurite density) correlate in all cases is unknown.

We find that the majority of significant causal effects are observed between pairs of brain phenotypes, which also exhibit large patterns of genetic correlation with each other. This result may be driven by the presence of latent causal factors which induce dependencies between groups of brain regions. Exploring this hypothesis further is an important consideration for future research.

We also observe that the effect of brain phenotypes on BD is consistently of larger magnitude than the effect of BD on brain phenotypes in both the TCE estimates and our dense DCE solution, consistent with previous results investigating the causal relationship between psychiatric conditions and brain imaging phenotypes^19^. This finding is interesting given that a degree of brain-state variation is plastic and can vary based on mood state, medication usage, or previous viral infection ^55–57^. Our results indicate that certain brain alterations, which can be characterized as brain-trait metrics, may precede BD onset, underscoring the importance of brain development in psychiatric pathology ^58^. The dysconnectivity hypothesis of BD neuroanatomy posits that aberrant connections between brain regions may give rise to the various cognitive features observed in individuals with BD ^59^. Our finding of white matter ICVF phenotypes as the main effectors of BD across TCE and DCE solutions may capture an aspect of this hypothesis, whereby less axonal tissue and decreased neurite density may impact information flow between indexed regions.

Aggregating phenotypic categories into ‘white matter’ or ‘non-white matter’, we find that white matter phenotypes across both DCE solutions have the largest out degree as measured by the number of non-zero outgoing edges. White matter tissue is comprised of more axonal fibers than gray matter, which by contrast is composed of more axonal endings and neuronal bodies. Larger effects of white matter phenotypes on BD implies that cognitive systems associated with BD are characterized by aberrant information transmission across and between brain regions. This effect is also observed at various levels of sparsity in output graphical lasso solutions that have accounted for the correlation structure of causal estimates between brain regions.

At the individual regions level, we find several results consistent with the previous literature. ICVF levels in the left anterior thalamic radiations was found to be a non-zero effector of BD in over 90% of stable output graph solutions in our work, with a previous study finding decreased FA in this fiber in individuals with BD ^60^. This tract connects nuclear groups in the thalamus to the frontal lobe, with the thalamus historically recognized as an important component of the limbic system ^61^. The limbic system is thought to play a key role in emotional processing tasks, especially in the context of BD; previous studies have hypothesized that dysfunction of limbic processes can contribute to BD symptoms ^10,62^. Another region of interest across solutions is the forceps minor, which is found in 50% of stable sparse output graph solutions, is an FDR-significant phenotype acting on BD, and is a non-zero effector of BD in our sparse DCE graph. The forceps minor is a fiber bundle connecting the lateral and medial surfaces of the frontal lobes and crosses the midline via the genu of the corpus callosum, and has been previously implicated in BD through decreased FA measures and reduced volume ^63–65^. Previous work has also identified shared loci between FA in the corpus callosum and BD ^66^. Other brain phenotypes of interest include diffusion measures in the superior and inferior longitudinal fasciculus, identified as non-zero effectors of BD across multiple sparse solutions (Figure S20). Our selected sparse DCE network implicated several measures of the ICVF in the superior longitudinal fasciculus, which is a large connection of associative fibers connecting frontal and anterior areas of the cerebrum. Bilateral tracts were also found to have lower FA in a previous mega-analysis of cohorts from multiple sites in the ENIGMA consortium ^53^.

Increased volumes of both the left and right caudate are FDR-significant effectors of BD according to *GSMR2* estimates. The same directions of effect are observed in our stable dense DCE solution and across MR methods. This is contrary to the direction of effect reported in a previous observational ENIGMA study ^53^. The caudate has been of historical interest in BD, with early neuroimaging studies describing increased activity in the left hemisphere ^67^. How volumetric changes in caudal structures (and the striatum complex to which they are related) may impact BD symptoms remains unknown, but previous studies have posited that its function is related to reward systems and memory ^68^. The striatum is also thought to be involved in addictive behaviors and associative learning, which are relevant cognitive systems related to BD pathology ^69^.

We found that only 7 of 28 FDR-significant BD-phenotype causal pairs have significant genetic correlation estimates. This indicates that conditioning phenotype inclusion for our MR analyses on the presence of significant genetic correlation estimates, as employed in previous studies ^19^, would miss relevant causal relationships, Generally, *rg* estimates and the causal relationships between those regions are not always consistent in direction or magnitude, as evidenced in our network diagrams. This implies that different information is captured by these distinct methods ^70^.

Finally, our predictive analysis attempts to ground our theoretical causal experiments in a practical application. While the variance explained on the liability scale is marginally higher using DCE weights compared to TCE weights in the Galway testing set, our limited sample size likely leaves us underpowered to detect any true effects or establish if either solution has any predictive capacity. Additionally, we observe the opposite effect in the Oslo population. To our knowledge, this marks the first application of MR for predictive applications at scale, which if realized, may have transformative benefits for the proactive treatment of psychiatric conditions. Future work could explore potential increases in predictive power by obtaining neuroimaging information on the most significant causal neuroimaging variables.

Some limitations of this study warrant noting. The sample size discrepancies between brain phenotypes and BD may impact the precision of our causal estimates. However, we have utilized the best-powered GWAS currently available for all phenotypes. Variants that are GWS for brain phenotype variation are measured in a population of individuals of primarily European ancestry aged 40-69, which may impact the generalizability of our results to other populations. We performed numerous tests to ensure the robustness of our MR results, in line with STROBE guidelines; however, it is difficult in practice to guarantee that all assumptions have been met. Future work to test the predictive capabilities of causal estimates in longitudinal BD neuroimaging samples would also be of great interest for future clinical applications. Finally, when testing for a difference in means between BD causal effects and causal effects on BD, accounting for the correlation between incoming and outgoing causal effects in similar phenotype categories may attenuate the significance of our independent t-test results. However, this is difficult to achieve in practice because each exposure is indexed by different instruments, thus making each test technically independent. At the category level, our application of a Bonferroni correction ensures that we are conservative in our estimation.

### Conclusions

Here, we leverage access to multiple well-powered GWAS to carry out a multitude of MR tests to generate new causal hypotheses while carrying out robust sensitivity analyses and sanity checks in the process. Our application of graphical lasso methods to causal estimates also offers interpretative benefit at both the individual component and systems level, providing direct causal estimates accounting for the causal relationships between brain regions. We find that white matter ICVF phenotypes and white matter phenotypes in general are consistent effectors of BD, implying that white matter microstructure disruptions have a causal relationship with BD. We also find that brain phenotype variation has larger effects on BD than *vice versa*, establishing a novel framework for conceptualizing BD pathology. We also attempt to establish that causal estimates can have interventional potential through predictive applications to separate datasets with limited success.

## Supplementary Information

Access to all main figures as high-resolution PDFs and the supplementary material is available here.

## Data availability

All summary statistics and software used in this analysis are publicly available. PGC3 BD summary statistics are available on the PGC download page (https://figshare.com/articles/dataset/PGC3_bipolar_disorder_GWAS_summary_statistics/14102594). UKBB neuroimaging summary statistics are available for download on the BIG40 website (https://open.win.ox.ac.uk/ukbiobank/big40/).

## Code availability

All code necessary to recreate this analysis are available at https://github.com/oconnell-s/causal_networks/.

## Acknowledgements

Research was conducted with the financial support of NIMH R01MH130879 “Delineating the network effects of mental disorder-associated variants using convex optimization methods” (PI David Knowles), grant number 18/CRT/6214 from Science Foundation Ireland, the Irish Research Council, the Health Research Board of Ireland (HRA-POR-324 Prof Cannon), NIH grant 1R01MH129742 - 01 to OAA., the Research council of Norway (#324499), and Nordforsk (#164218).

## Conflicts of Interest

Dr. Andreassen has received speaker fees from Lundbeck, Janssen, Otsuka, and Sunovion and is a consultant to Cortechs.ai. and Precision Health. Dr. Westlye is a shareholder of baba.vision. All other authors report no competing interests.

## Notes

### Author Declarations

The use of the Norwegian Thematically Organized Psychosis (TOP) sample data was approved by the Regional Committee for Medical Research Ethics. Ethical approval was received by the University College Hospital Galway Research Ethics Committee, and participants gave written fully informed consent before participating.

