## Supplementary Materials for "Deriving Mendelian Randomization-based Causal Networks of Brain Imaging Phenotypes and Bipolar Disorder"

### Supplemental Note: Deriving Causal Networks of Brain Imaging Phenotypes and Bipolar Disorder Using Mendelian Randomization and Inverse Sparse Regression

Shane O'Connell

December 2024

### 1 Supplementary figures

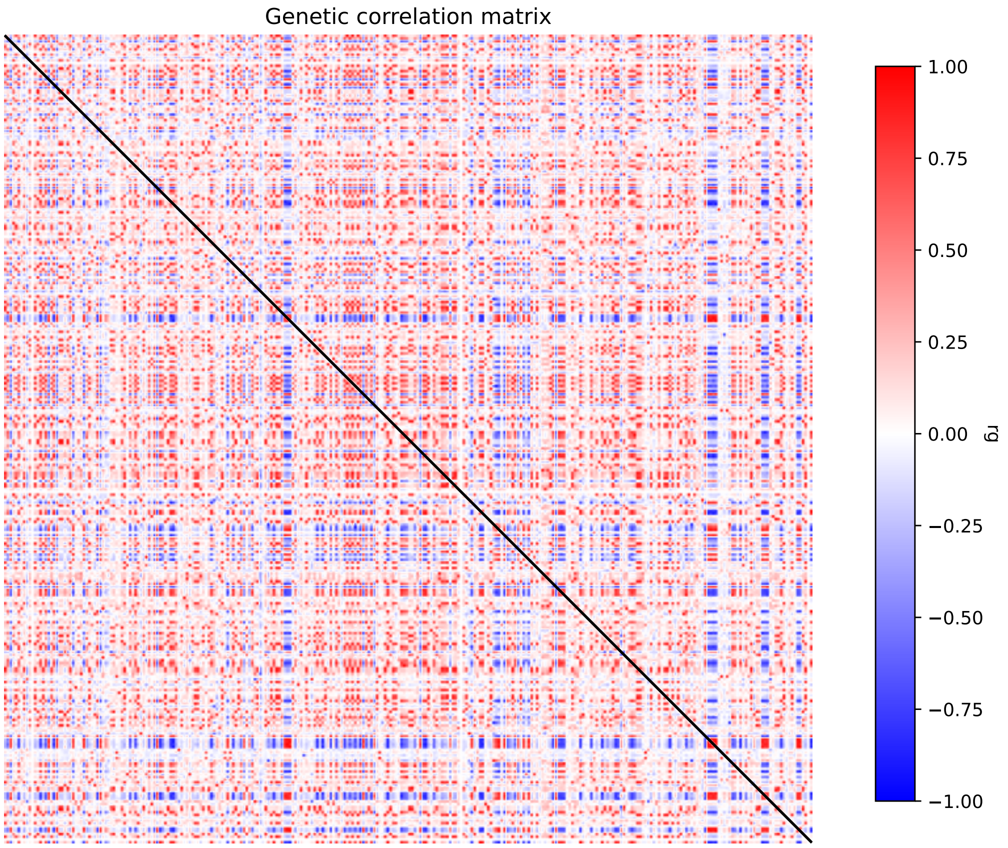

Figure S1: Genetic correlation ($rg$) matrix of every pair of phenotypes in the network calculated using *ldsc*.

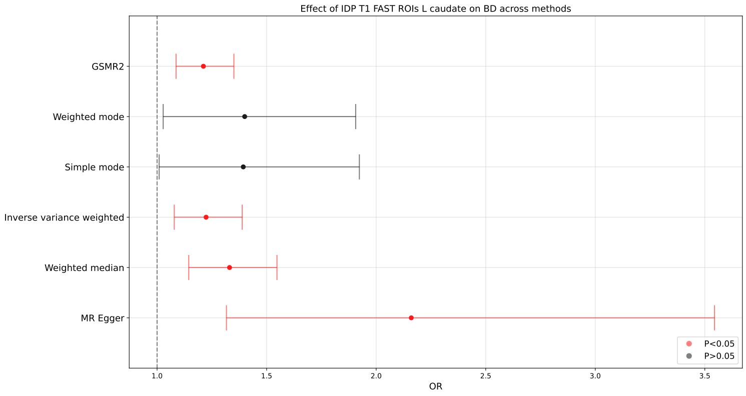

(a)

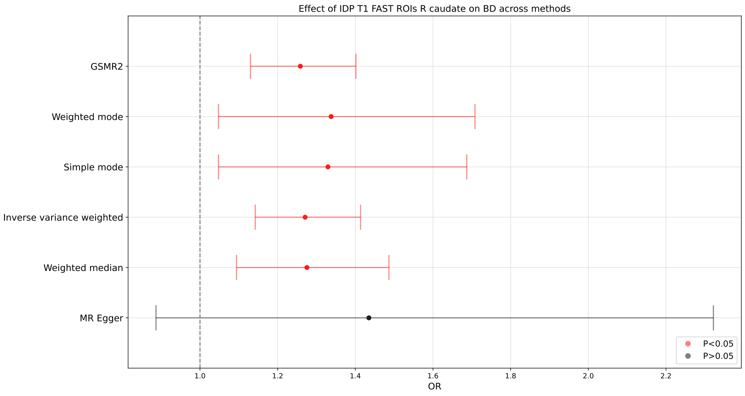

(b)

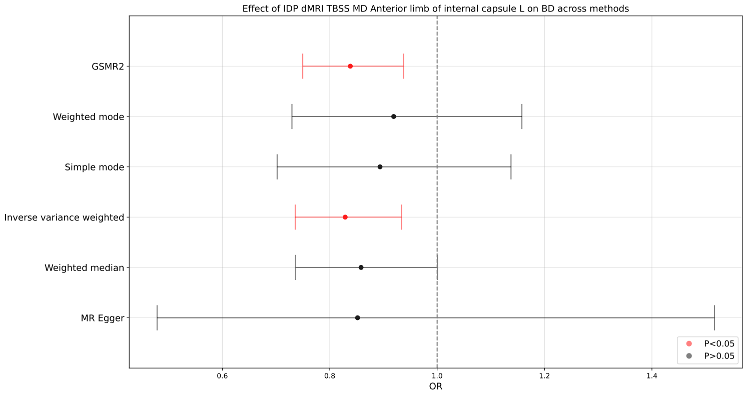

(c)

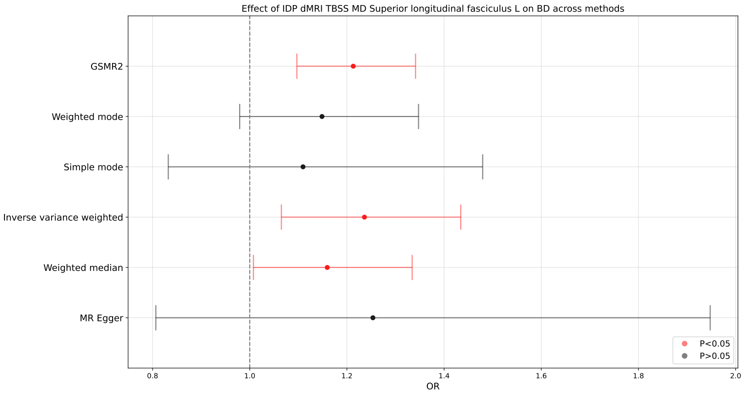

(d)

Figure S2: Plot of estimated causal effect of IDP 124 (a), 125 (b), 1619 (c), and 1643 (d) on BD across 6 different MR methods (GSMR2, Weighted mode, Simple mode, inverse variance weighted, Weighted median, and MR Egger).

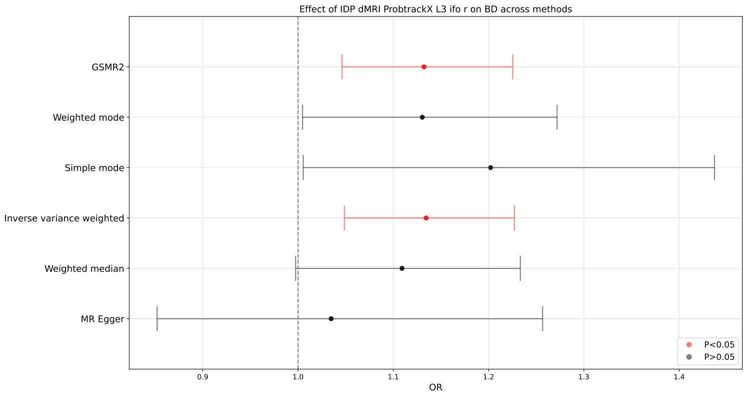

(a)

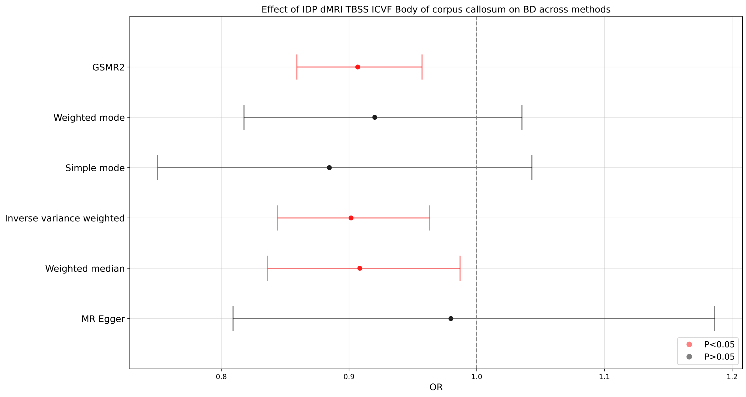

(b)

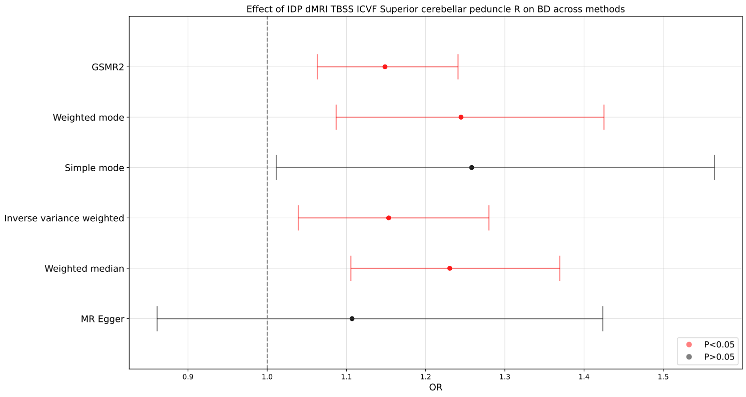

(c)

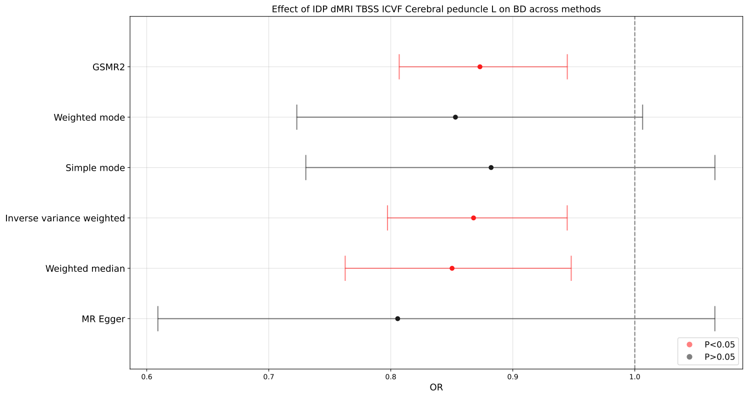

(d)

Figure S3: Plot of estimated causal effect of IDP 1888 (a), 1905 (b), 1914 (c), and 1917 (d) on BD across 6 different MR methods (GSMR2, Weighted mode, Simple mode, inverse variance weighted, Weighted median, and MR Egger).

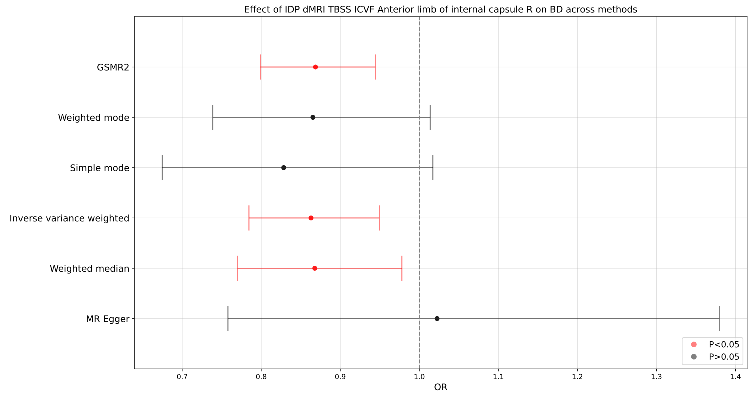

(a)

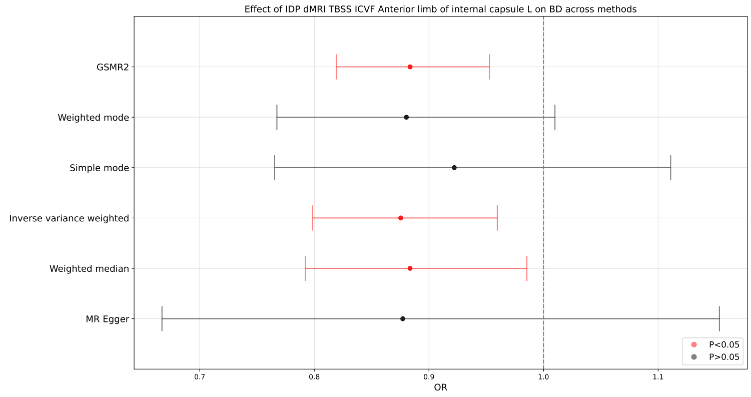

(b)

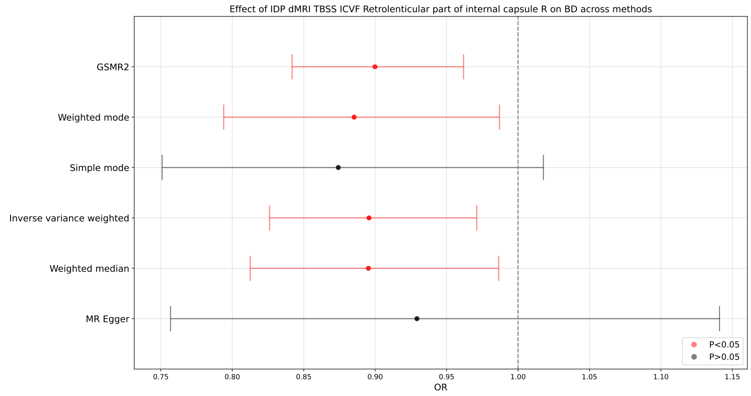

(c)

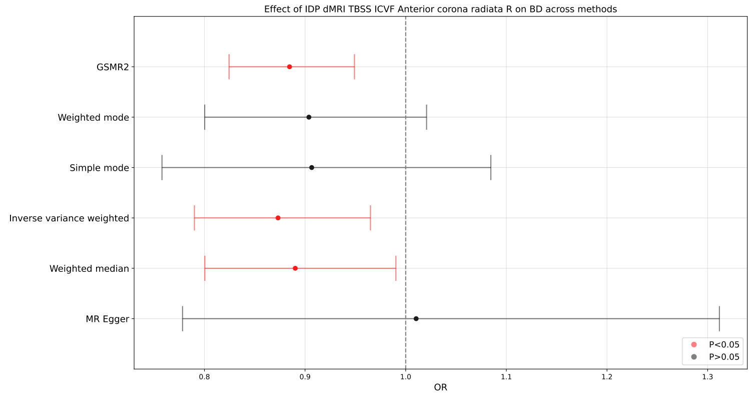

(d)

Figure S4: Plot of estimated causal effect of IDP 1918 (a), 1919 (b), 1922 (c), and 1924 (d) on BD across 6 different MR methods (GSMR2, Weighted mode, Simple mode, inverse variance weighted, Weighted median, and MR Egger).

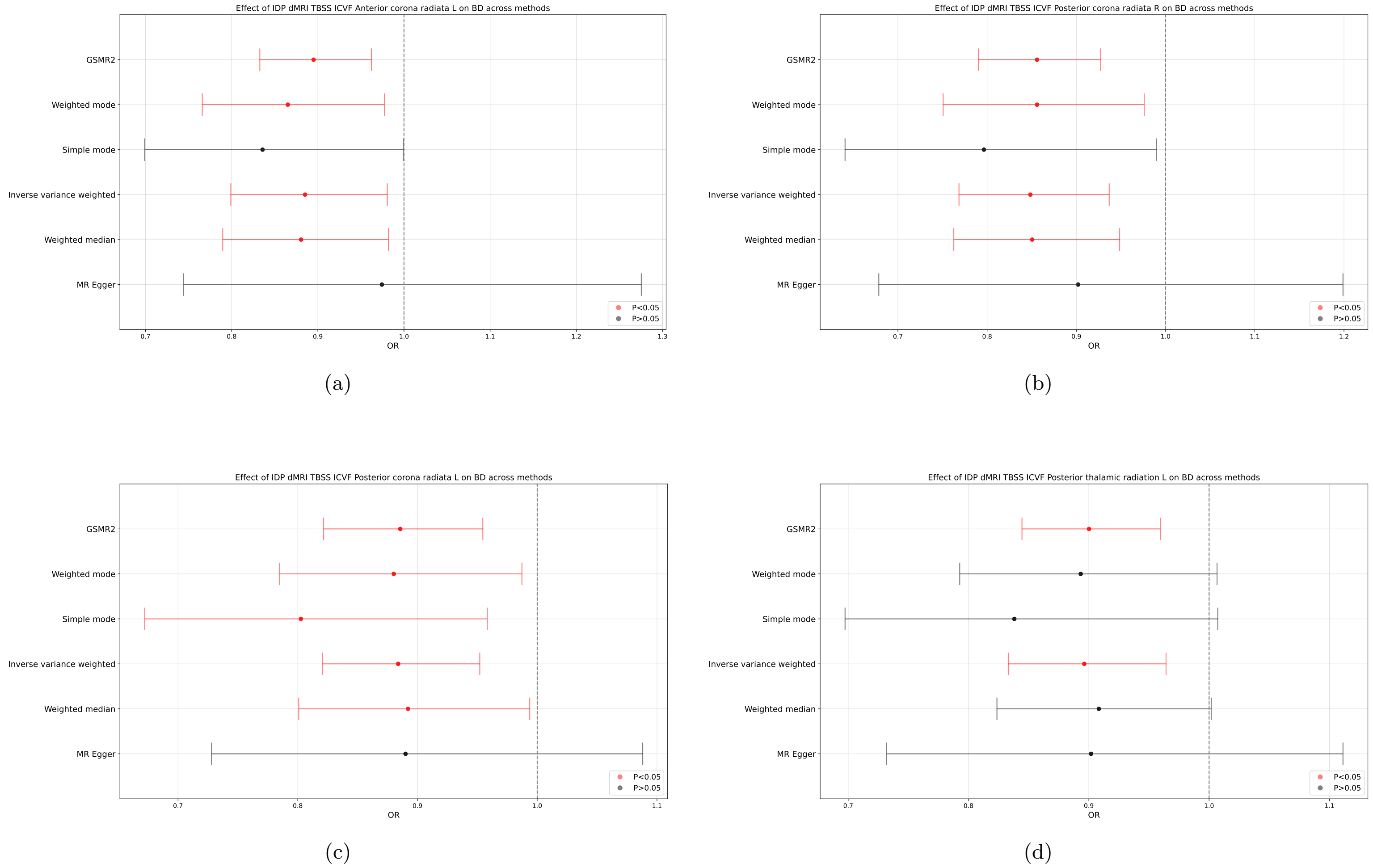

Figure S5: Plot of estimated causal effect of IDP 1925 (a), 1928 (b), 1929 (c), and 1931 (d) on BD across 6 different MR methods (GSMR2, Weighted mode, Simple mode, inverse variance weighted, Weighted median, and MR Egger).

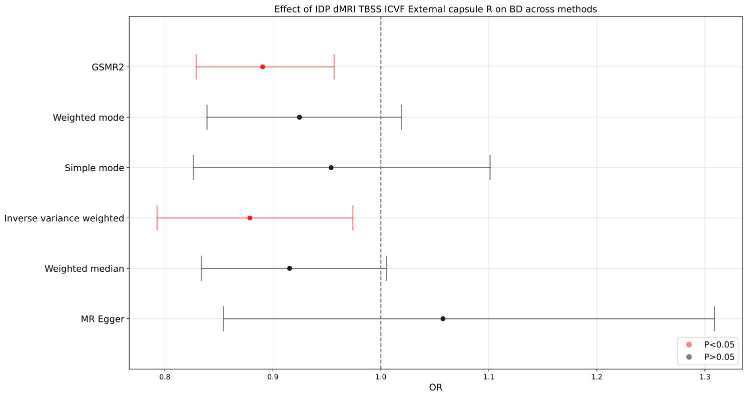

(a)

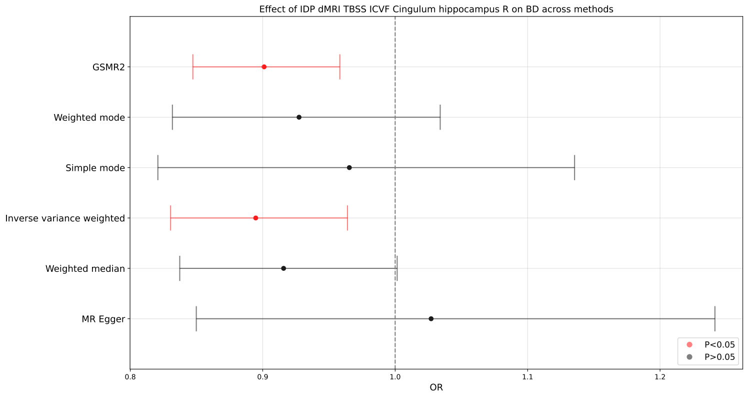

(b)

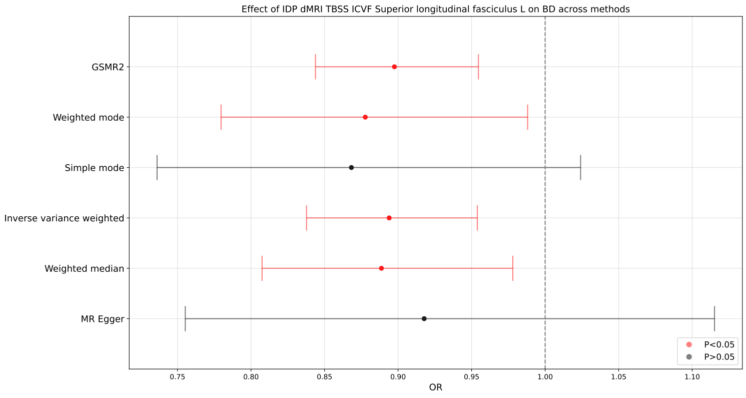

(c)

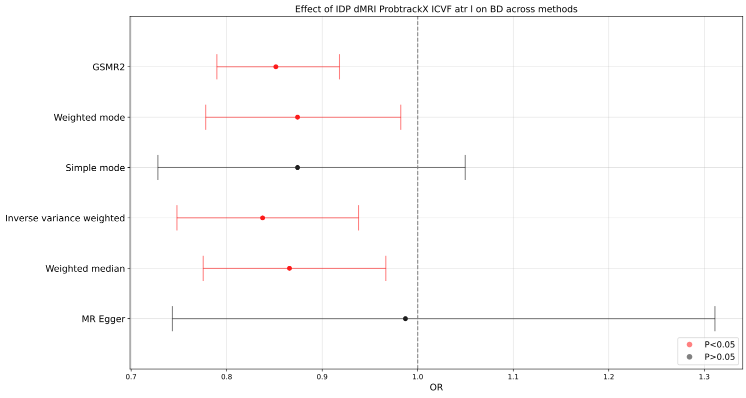

(d)

Figure S6: Plot of estimated causal effect of IDP 1934 (a), 1938 (b), 1943 (c), and 1952 (d) on BD across 6 different MR methods (GSMR2, Weighted mode, Simple mode, inverse variance weighted, Weighted median, and MR Egger).

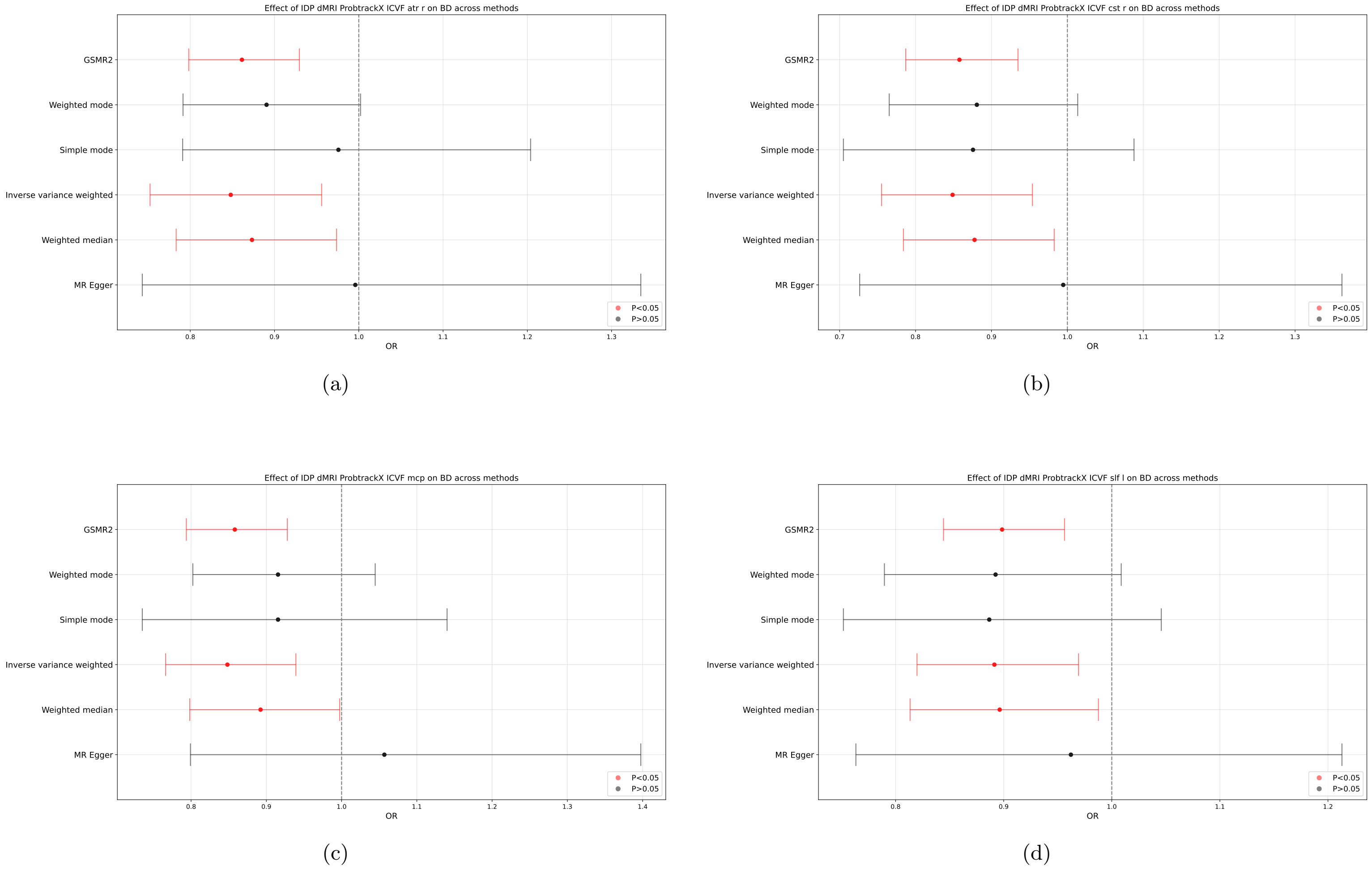

Figure S7: Plot of estimated causal effect of IDP 1953 (a), 1959 (b), 1966 (c), and 1971 (d) on BD across 6 different MR methods (GSMR2, Weighted mode, Simple mode, inverse variance weighted, Weighted median, and MR Egger).

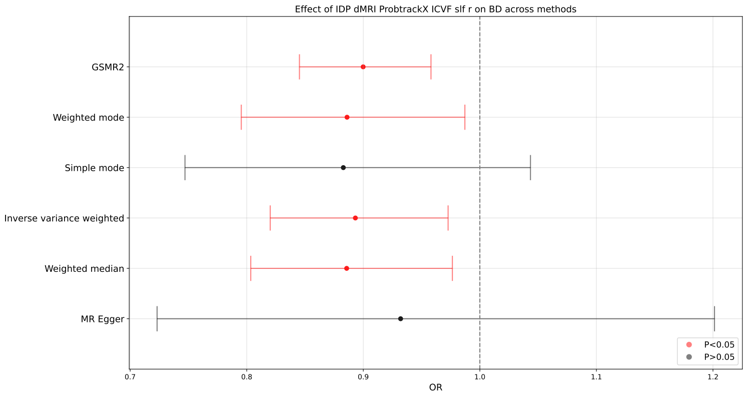

(a)

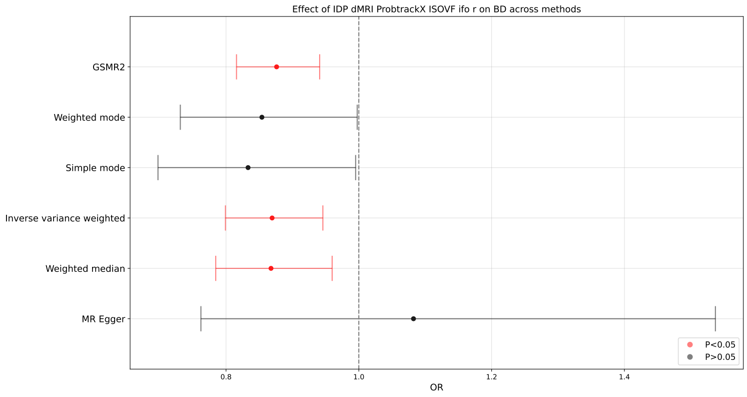

(b)

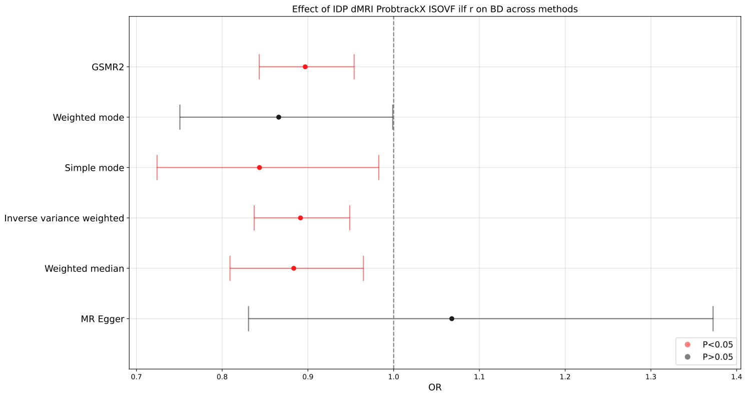

(c)

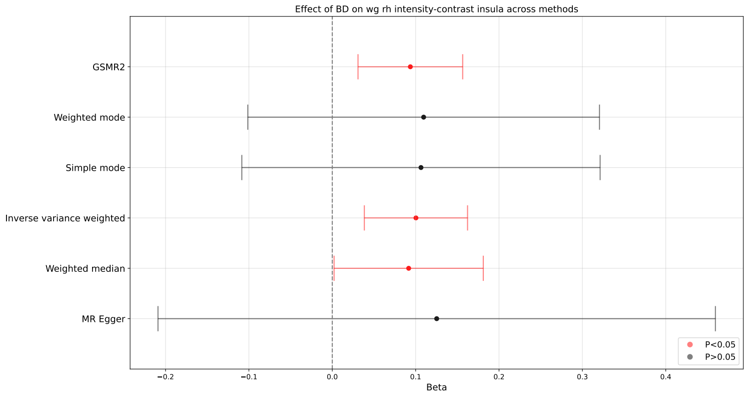

(d)

Figure S8: Plot of estimated causal effect of IDP 1972 (a), 2113 (b), 2115 (c) on BD, and BD on IDP 1436 (d) across 6 different MR methods (GSMR2, Weighted mode, Simple mode, inverse variance weighted, Weighted median, and MR Egger).

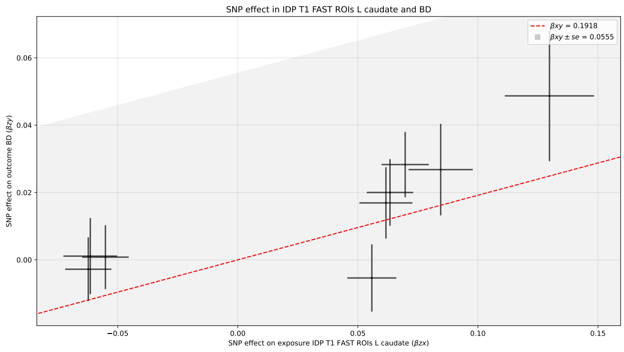

(a)

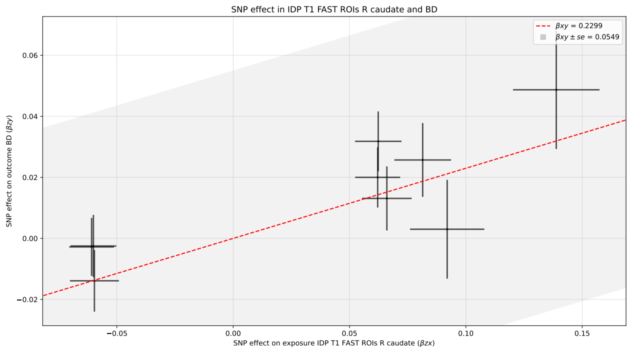

(b)

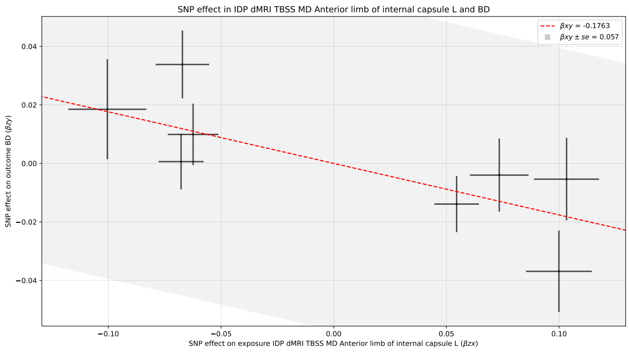

(c)

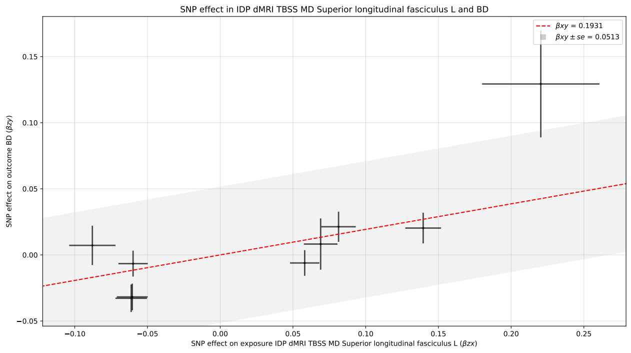

(d)

Figure S9: Variant-exposure variant-outcome plot of SNP instruments describing the estimated causal effect of IDP 124 (a), 125 (b), 1619 (c), and 1643 (d) on BD.

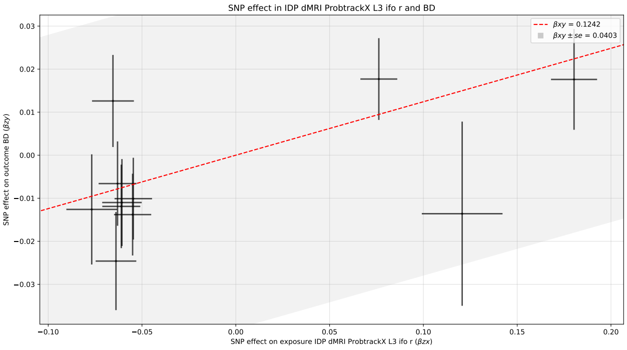

(a)

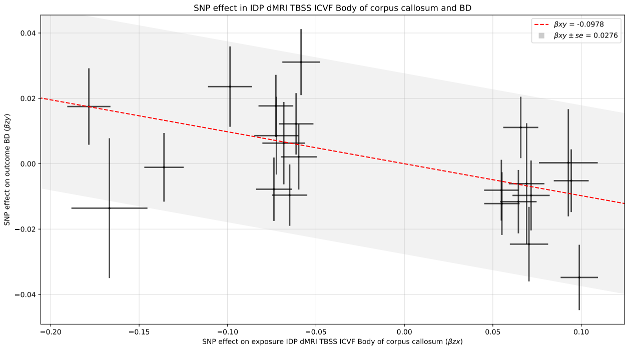

(b)

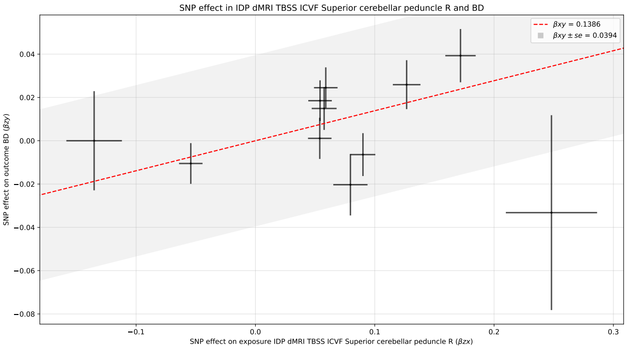

(c)

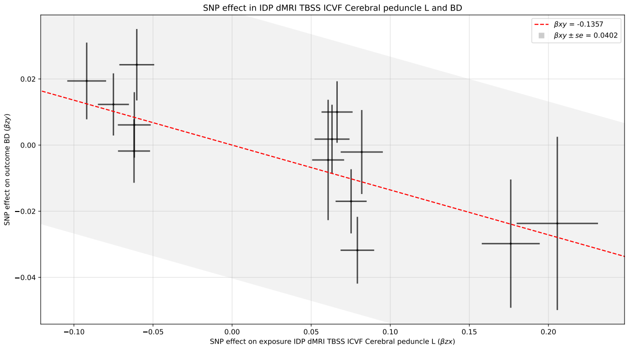

(d)

Figure S10: Variant-exposure variant-outcome plot of SNP instruments describing the estimated causal effect of IDP 1888 (a), 1905 (b), 1914 (c), and 1917 (d) on BD.

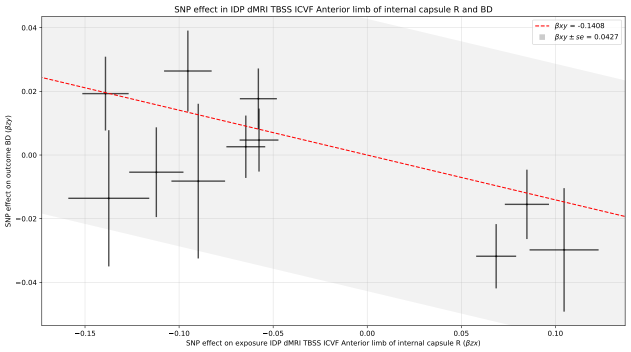

(a)

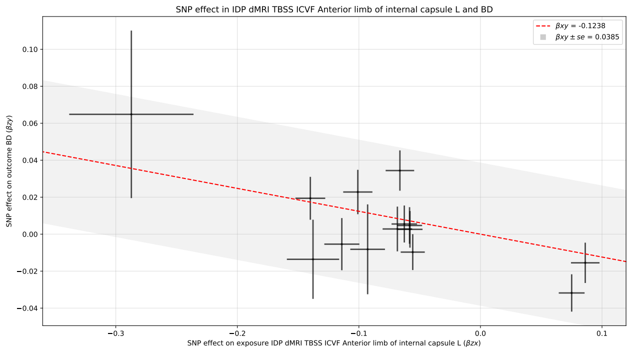

(b)

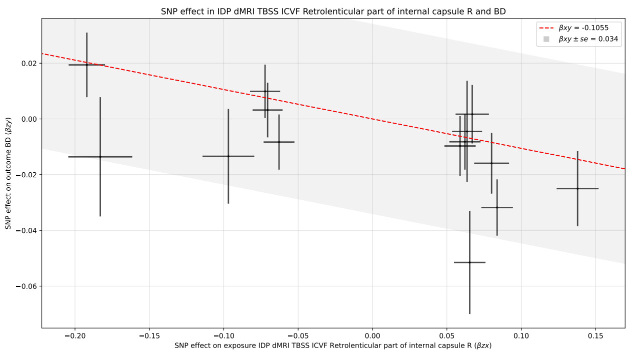

(c)

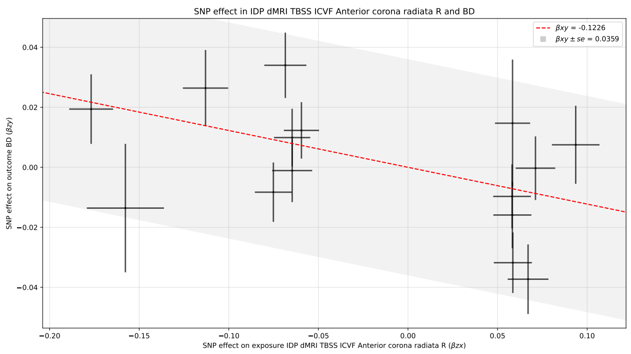

(d)

Figure S11: Variant-exposure variant-outcome plot of SNP instruments describing the estimated causal effect of IDP 1918 (a), 1919 (b), 1922 (c), and 1924 (d) on BD.

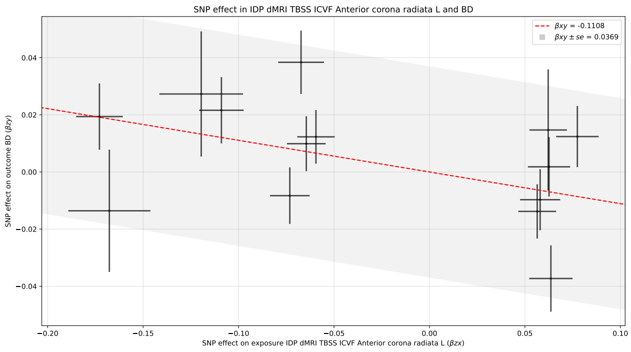

(a)

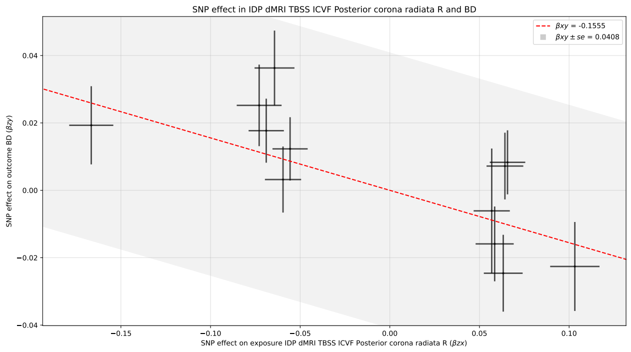

(b)

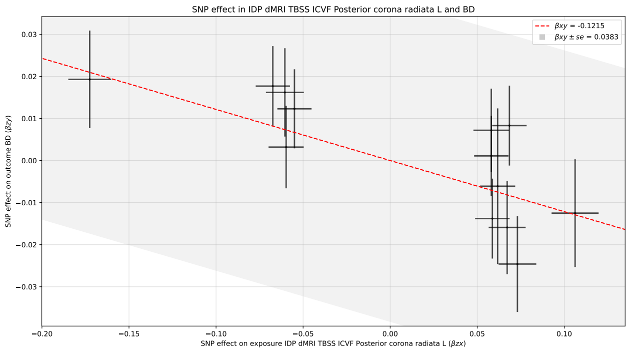

(c)

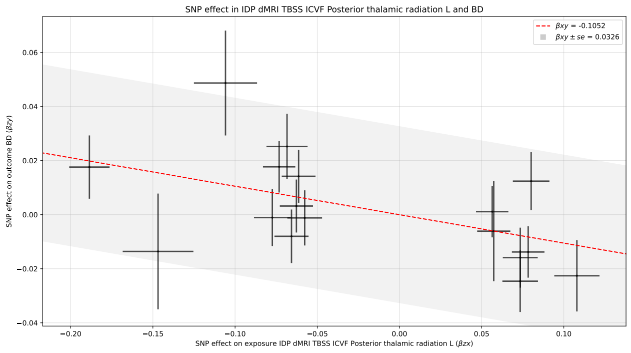

(d)

Figure S12: Variant-exposure variant-outcome plot of SNP instruments describing the estimated causal effect of IDP 1925 (a), 1928 (b), 1929 (c), and 1931 (d) on BD.

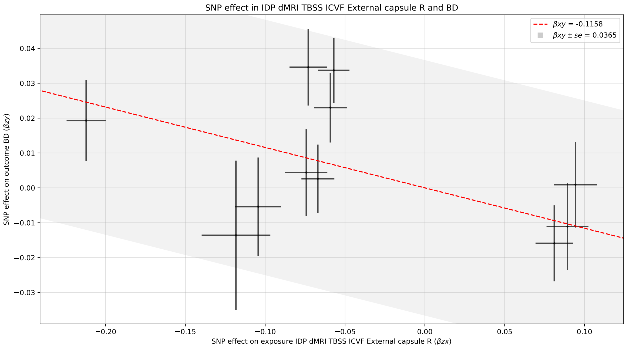

(a)

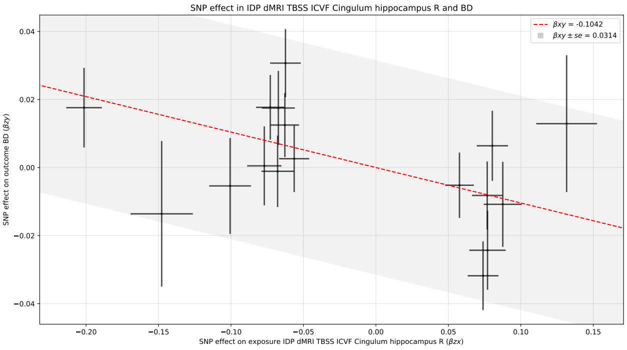

(b)

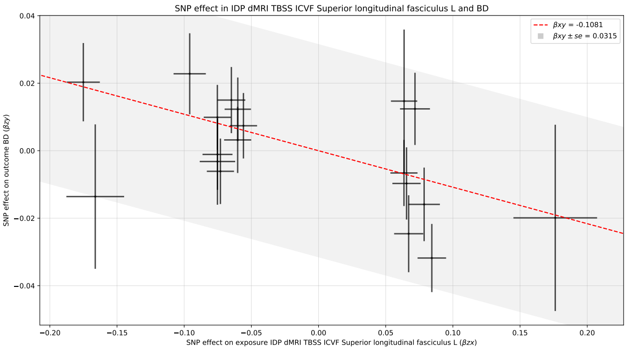

(c)

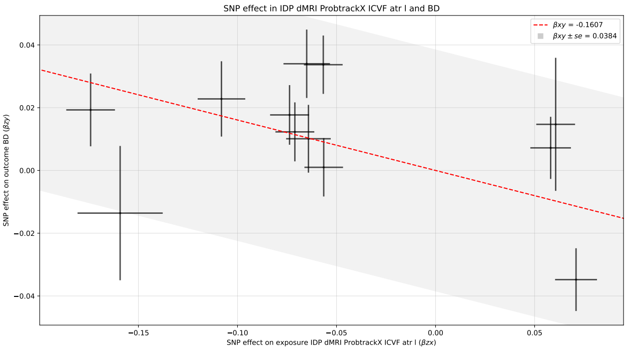

(d)

Figure S13: Variant-exposure variant-outcome plot of SNP instruments describing the estimated causal effect of IDP 1934 (a), 1938 (b), 1943 (c), and 1952 (d) on BD.

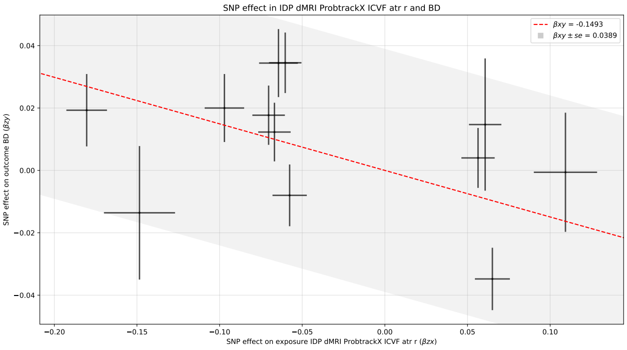

(a)

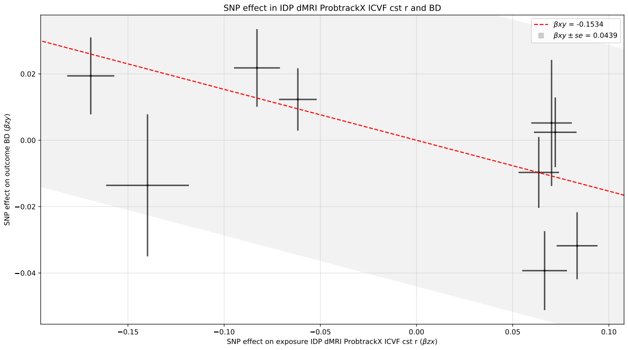

(b)

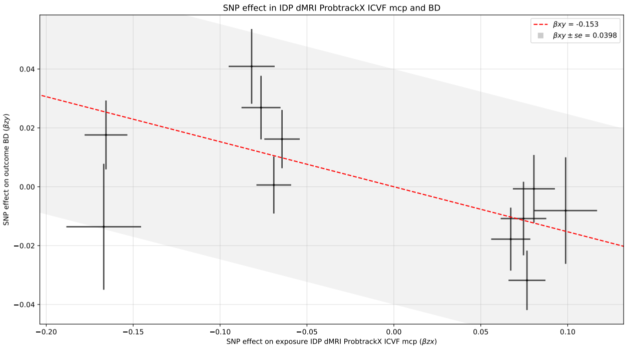

(c)

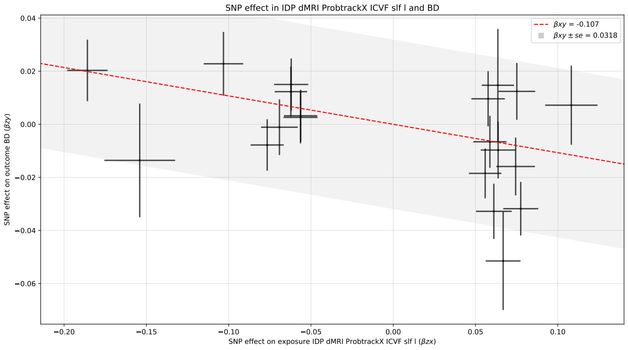

(d)

Figure S14: Variant-exposure variant-outcome plot of SNP instruments describing the estimated causal effect of IDP 1953 (a), 1959 (b), 1966 (c), and 1971 (d) on BD.

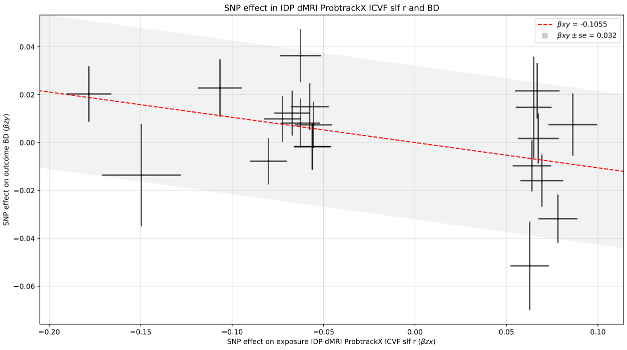

(a)

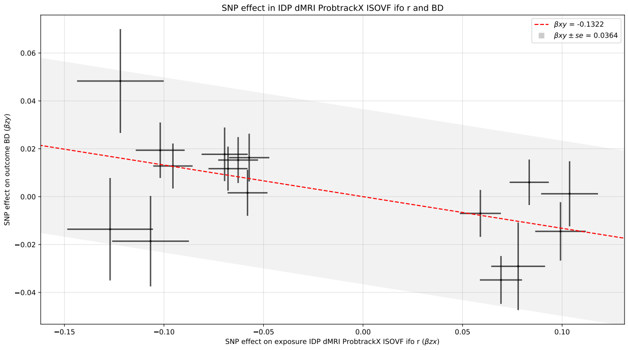

(b)

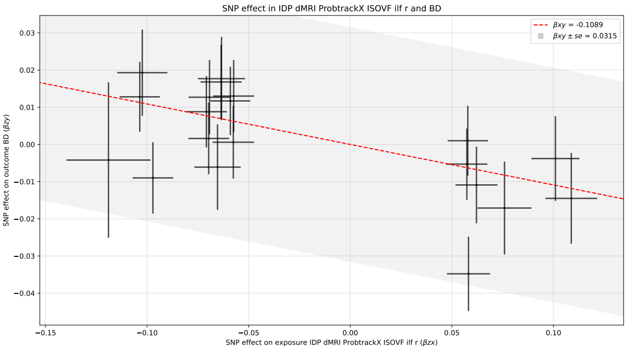

(c)

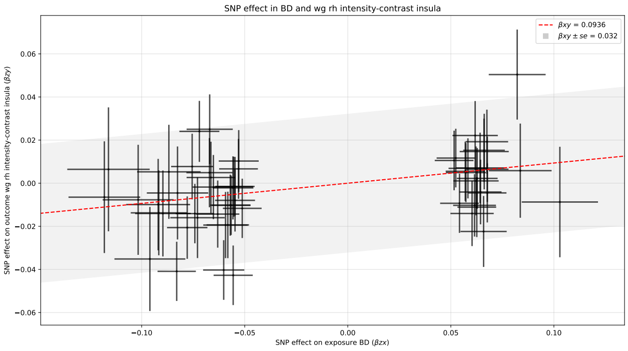

(d)

Figure S15: Variant-exposure variant-outcome plot of SNP instruments describing the estimated causal effect of IDP 1972 (a), 2113 (b), 2115 (c) on BD, and BD on IDP 1436 (d).

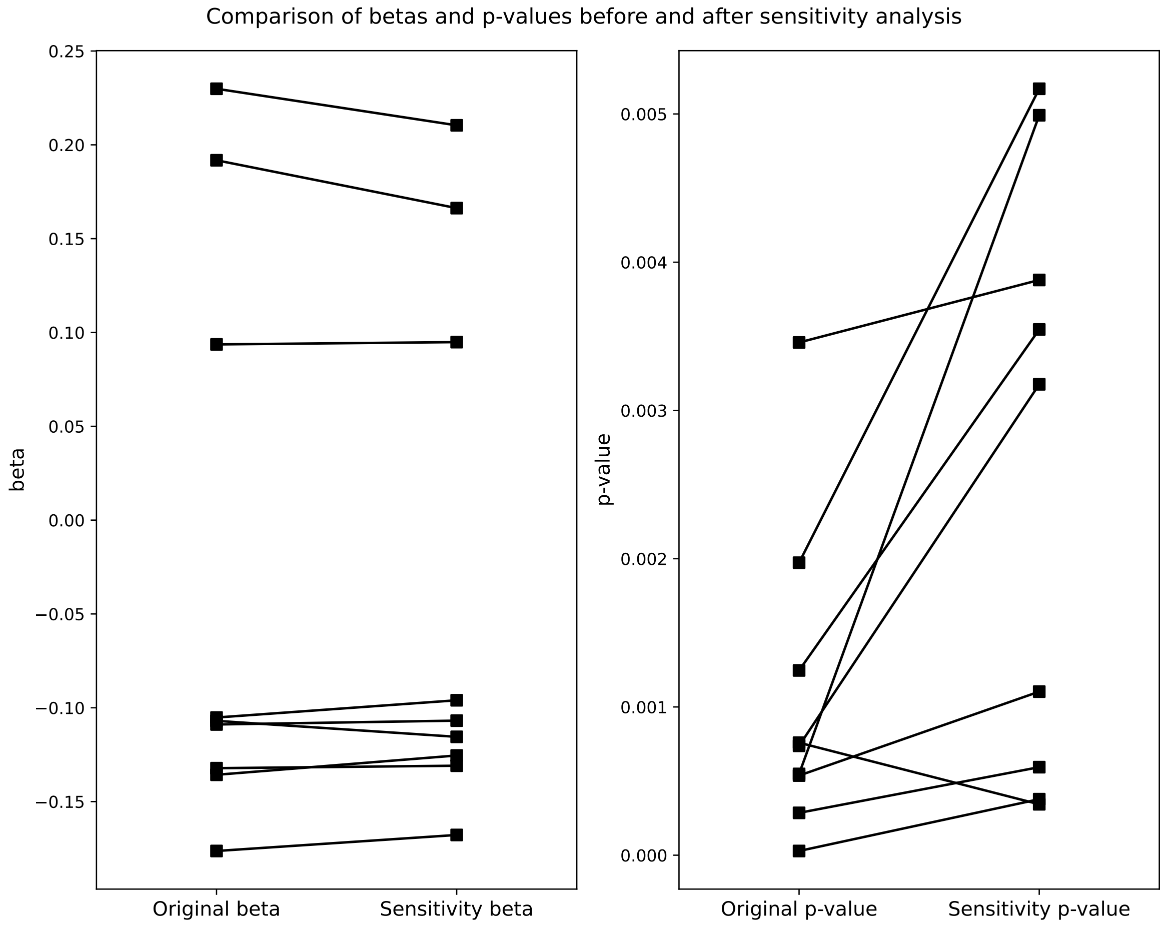

Figure S17: Dotplots of beta (left) and p-value (right) changes across tests before and after confounder-associated SNP exclusion. The original column (left of each panel) denotes the test results with all instruments included, and the right column (right of each panel) denotes the test results after instruments associated with a panel of confounder phenotypes (described in Methods section) were removed. All relevant tests remained significant after SNP exclusion.

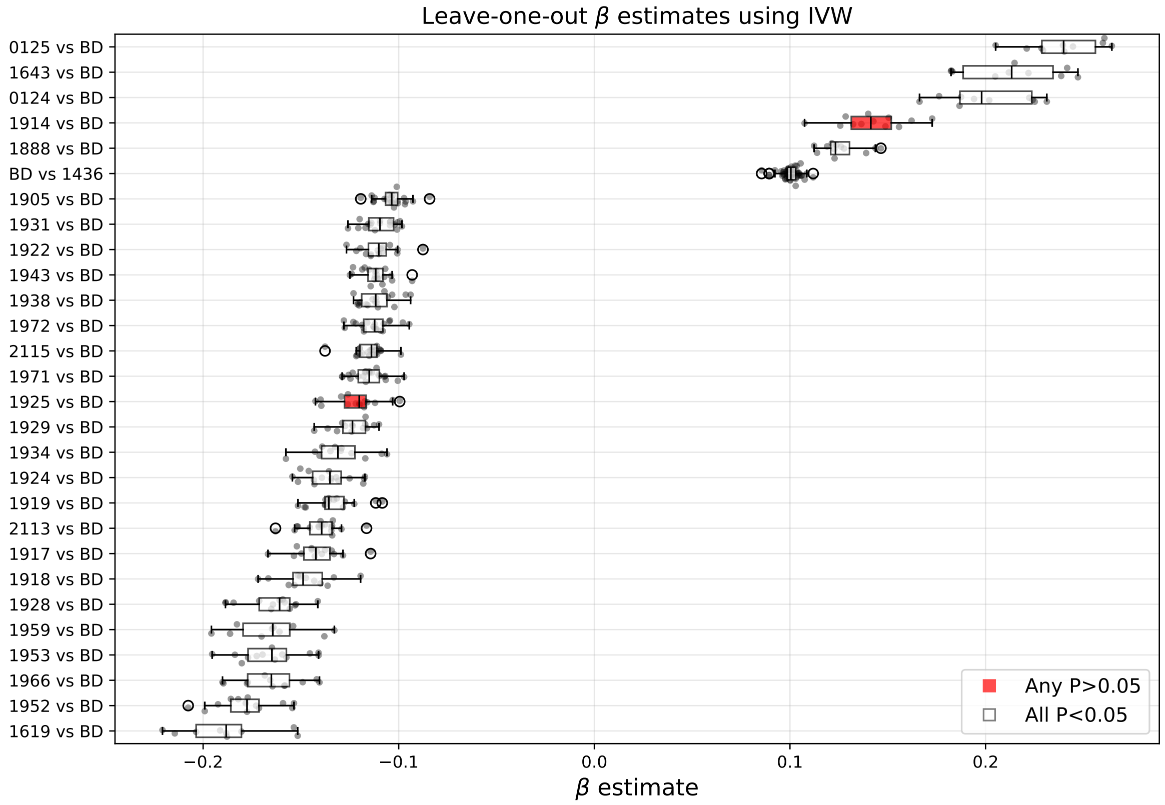

Figure S16: Boxplots of leave-one-out analyses across 28 FDR-significant MR pairs. Beta coefficients from an inverse-variance weighted regression are presented per pair, whereby each point represents a test with one instrument excluded per valid instrument. Where any test statistics lost significance after instrument removal, the boxplot was colored red.

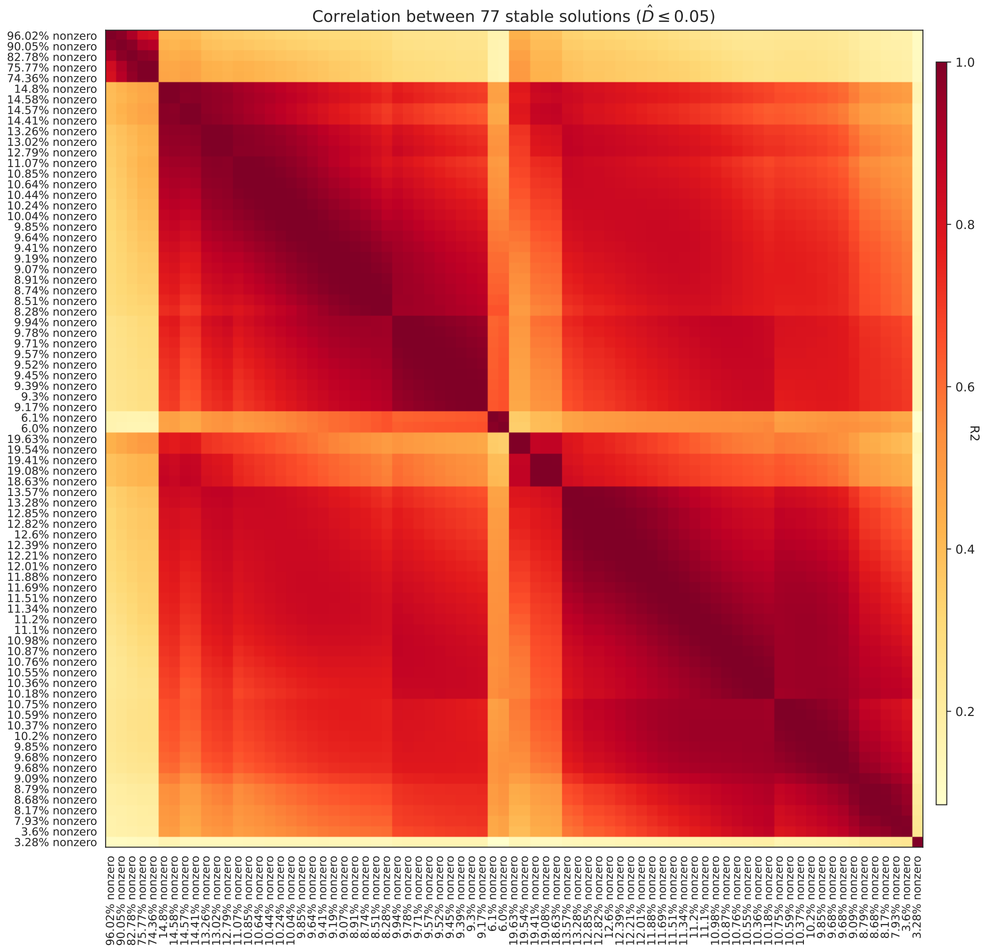

Figure S18: Correlation matrix of 77 stable output direct causal effect (DCE) solutions. Stable solutions were determined by the value of $\hat{D}$, whereby solutions with values less than 0.05 had high probabilities of replication across random graph re-samplings. Further details on $\hat{D}$ calculation can be found in the main text and in [1],[2].

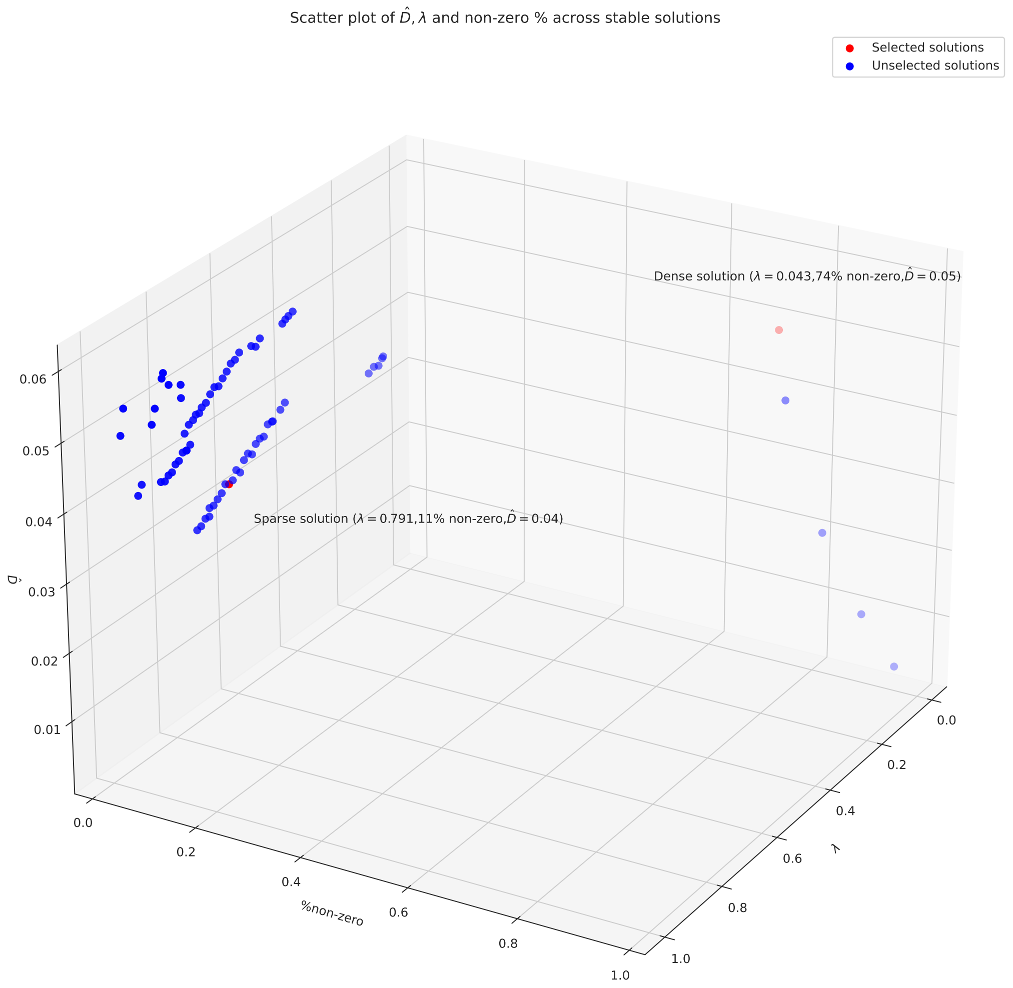

Figure S19: Scatter plot of $\hat{D}$ (stability metric) vs. $\lambda$ (regularization strength) vs. % of non-zero entries in output matrix across 77 stable solutions presented in Figure S18. Solutions analysed in the main text are highlighted in red.

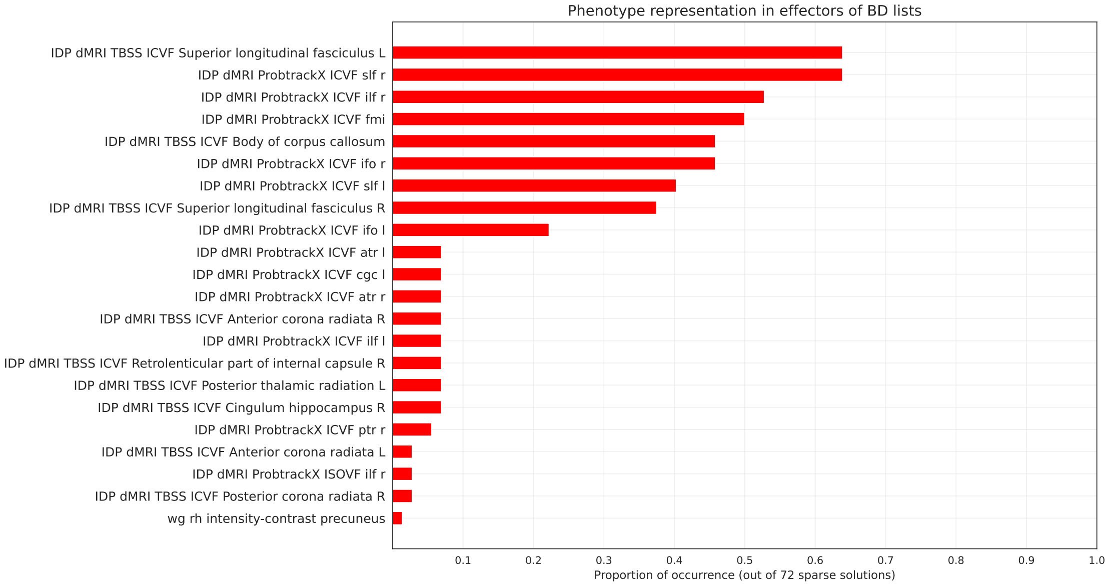

Figure S20: Barplot of phenotype occurrence as non-zero effector of BD across 72 sparse solutions (maximum non-zero % $\approx$ 20). Phenotype names are listed along the y-axis and proportion of occurrence as a fraction of the total number of solutions is presented along the x-axis.

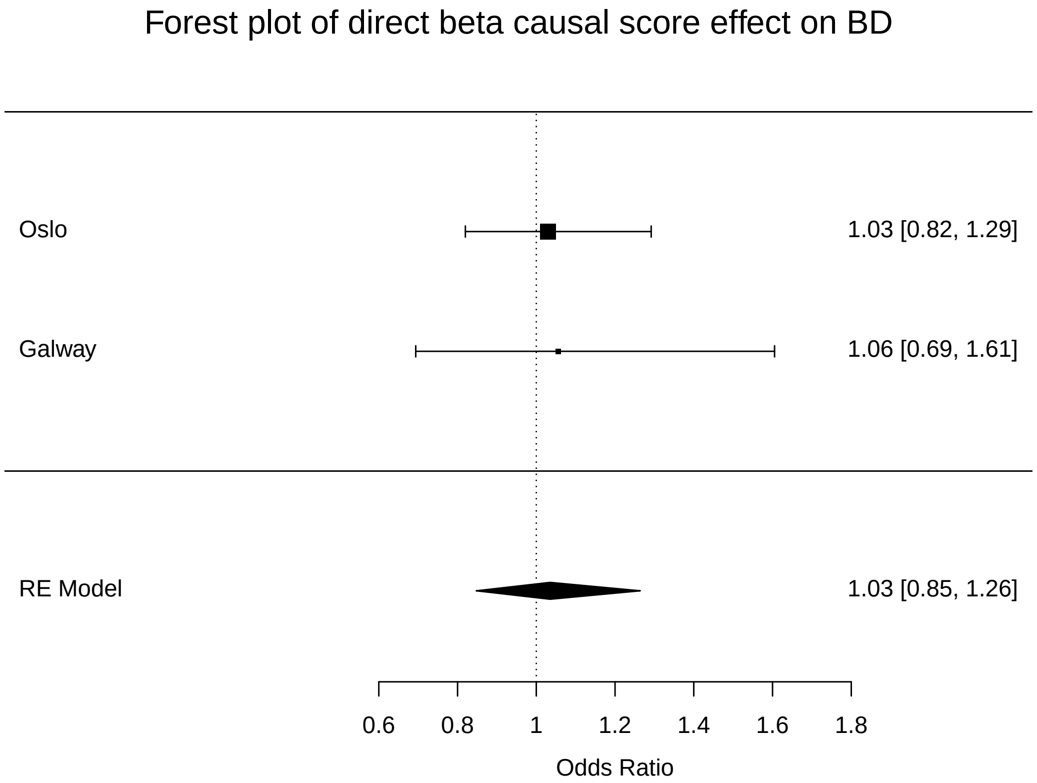

Figure S21: Forest plot of BD status regressed against direct $\beta$ causal score in two cohorts, named Oslo and Galway, in the odds ratio scale. The bottom row describes a random effects meta-analysis, carried out using the *metafor* R package.

#### 2 Supplementary tables

| Phenotype Category | Count in network | Median $h^2$ | $N$ of instruments $\mu$ ($\sigma$) |
|---|---|---|---|
| regional and tissue volume | 71 | 0.2865 | 15.70 (12.75) |
| WM tract ICVF | 61 | 0.3230 | 17.49 (3.71) |
| cortical grey-white contrast | 57 | 0.3301 | 19.67 (5.70) |
| WM tract diffusivity | 37 | 0.2796 | 11.97 (1.71) |
| cortical area | 28 | 0.2751 | 15.61 (4.95) |
| WM tract OD | 9 | 0.2858 | 11.11 (1.05) |
| regional and tissue intensity | 8 | 0.2176 | 33.50 (38.12) |
| WM tract FA | 8 | 0.2907 | 11.38 (1.69) |
| WM tract ISOVF | 6 | 0.2394 | 16.50 (3.78) |
| regional T2* | 6 | 0.2712 | 23.00 (7.48) |
| rfMRI connectivity | 3 | 0.3097 | 12.00 (1.00) |
| WM tract MO | 2 | 0.2612 | 14.50 (3.54) |
| white matter hyperintensity volume | 1 | 0.2580 | N/A |

Table S1: Cateogry counts, median heritabilities, and mean/s.d. ($\mu$ ($\sigma$)) number of instruments in 297 imaging derived phenotypes included in analyses. All phenotypes $\geq$ 10 genome-wide significant instruments based on clumping using the HRC reference panel and the following parameters: LD $r^2$ > 0.001, window 10,000 kb, $P \leq 5e^{-8}$.

| | **Oslo** | **Galway** |
|---|---|---|
| **Total N** | 565 | 100 |
| **Sex** | Male=269, Female=296 | Male=46, Female=54 |
| **Mean Age (s.d.)** | 32.64 (11.5) | 41.9 (13.05) |
| **N cases** | 127 (44 Male, 83 Female) | 44 (22 Male, 22 Female) |
| **Mean Age by diagnosis (s.d.)** | BD=31 (10.1), Control=33.1 (11.8) | BD=43.5 (12.4), Control=40.61 (13.5) |

Table S2: Demographic information of cohorts used for the application of direct causal $\beta$ score approach. Full sample acquisition information is detailed below.

### 3 Supplementary Note

#### 3.1 *inspre* execution details

The 77 stable solutions presented in the main text resulted from two methodological approaches which require discussion of the algorithm. The calculation of graph output stability is derived from the number of non-zero edges present across random re-samplings of the input graph at a particular $\lambda$ value. The proportion of times a non-zero edge is present is used to calculate $\hat{D}$, which is explained in greater detail in [1] and [2]. Because $\hat{D}$ was designed to measure the stability of sparse graphs, usually a user-specified threshold is imposed to avoid small values from being counted as non-zero edges. This value by default is set to $1 \times 10^{-5}$, which resulted in 38 solutions with $\hat{D}$ values at or below 0.05. This produced our denser solutions (the 5 in the right hand corner of Figure S19). We re-ran our analysis with a more stringent minimum value ($1 \times 10^{-3}$) to obtain a sparser family of graph solutions, resulting in 39 stable solutions which are mostly clustered in the left corner of Figure S19. We chose the dense solution with the smallest non-zero value percentage from our first method iteration and a sparse solution from the second method iteration. As can be seen from Figure S18, the correlation between solutions was generally high (median $\rho = 0.76$), and solutions of similar numerical density had higher concordance.

#### 3.2 Oslo cohort information

##### 3.2.1 Scanner

We use a General Electric Discovery MR750 3T scanner with a 32-channel head coil. With a spin echo planar imaging (EPI) sequence, the following parameters were used: repetition time (TR) of 8150 ms, echo time (TE) of 83.1 ms, and flip angle (FA) of 90°, field of view (FOV) of 256 × 256 mm, slice thickness of 2 mm, and an in-plane resolution of 2 mm. For diffusion weighted data, 10 volumes of b = 0, 60 of b = 1000s/mm2, and 30 of b = 2000s/mm2 were acquired. An additional 7 volumes of b = 0 with reversed phase-encoding direction were acquired for susceptibility distortion correction.

##### 3.2.2 Ethics

The use of the Norwegian Thematically Organized Psychosis (TOP) sample data was approved by the Regional Committee for Medical Research Ethics.

#### 3.3 Galway cohort information

##### 3.3.1 Scanner

MRI data were obtained on a 3T Achieva scanner (Philips, Best, The Netherlands) at the Welcome Trust Health Research Board National Centre for Advanced Medical Imaging at St. James's Hospital Dublin,

Ireland. High-resolution 3-dimensional T1-weighted turbo field echo magnetizationprepared rapid gradient-echo sequence was acquired using an 8-channel head coil (repetition/echo times = 8.5/3.046 ms, 1mm3 voxel size). Diffusion-weighted images were acquired at b = 1200 s/mm2 along with a single nondiffusion-weighted image (b = 0), using high angular resolution diffusion imaging involving 61 diffusion gradient directions, 1.8 3 1.8 3 1.9 mm voxel dimension, and field of view 198 3 259 3 125 mm.

##### 3.3.2 Ethics

Ethical approval was received by the University College Hospital Galway Research Ethics Committee, and participants gave written fully informed consent before participating

#### 3.4 Interactive networks download and instructions

Full networks can be downloaded from the following URL. To open the page,

- Unzip the archive *networks_app.zip*
- This should create a folder called *networks_app*.
- Navigate to the folder (using finder on Mac or any other file navigation tool)
- Open the *index.html* file
- Your networks in browser should render successfully.

Any issues, please feel free to email me here.
