## Supplementary material for "Deriving Mendelian Randomization-based Causal Networks of Brain Imaging Phenotypes and Bipolar Disorder": Table 1

| Exposure | Outcome | Beta/OR | 95% C.I. [lower, upper] | P | N snps | *rg* | P(rg) | *h2 (exposure)* | DCE beta (dense) | DCE beta (sparse) | UKBID | Any P > 0.05 (LOO) | N significant methods | Phenotype category |
|---|---|---|---|---|---|---|---|---|---|---|---|---|---|---|
| IDP dMRI TBSS ICVF Posterior corona radiata L | BD | 0.8856 | [0.8216, 0.9546] | 1.497E-03 | 13 | -0.0916 | 1.429E-02 | 0.3286 | -0.0059 | 0 | 1929 | FALSE | 5 | WM tract ICVF |
| IDP T1 FAST ROIs R caudate | BD | 1.2584 | [1.13, 1.4015] | 2.843E-05 | 9 | 0.0705 | 7.625E-02 | 0.2764 | 0.0299 | 0 | 125 | FALSE | 5 | regional and tissue volume |
| IDP dMRI ProbtrackX ICVF slf r | BD | 0.8999 | [0.8452, 0.9581] | 9.687E-04 | 20 | -0.0365 | 2.894E-01 | 0.3771 | -0.002 | -0.0047 | 1972 | FALSE | 4 | WM tract ICVF |
| IDP dMRI ProbtrackX ISOVF ilf r | BD | 0.8968 | [0.8432, 0.9539] | 5.373E-04 | 20 | -0.0146 | 6.974E-01 | 0.2462 | -0.013 | 0 | 2115 | FALSE | 4 | WM tract ISOVF |
| IDP dMRI TBSS ICVF Superior longitudinal fasciculus L | BD | 0.8975 | [0.8438, 0.9546] | 5.916E-04 | 19 | -0.0599 | 8.684E-02 | 0.3536 | -0.0026 | -0.0057 | 1943 | FALSE | 4 | WM tract ICVF |
| IDP dMRI TBSS ICVF Retrolenticular part of internal capsule R | BD | 0.8999 | [0.8418, 0.9619] | 1.913E-03 | 14 | -0.0499 | 1.802E-01 | 0.3163 | -0.0006 | 0 | 1922 | FALSE | 4 | WM tract ICVF |
| IDP dMRI TBSS ICVF Anterior corona radiata L | BD | 0.8951 | [0.8327, 0.9622] | 2.641E-03 | 14 | -0.0718 | 3.183E-02 | 0.3566 | -0.0016 | 0 | 1925 | TRUE | 4 | WM tract ICVF |
| IDP dMRI TBSS ICVF Posterior corona radiata R | BD | 0.856 | [0.7902, 0.9272] | 1.385E-04 | 12 | -0.0813 | 2.899E-02 | 0.3287 | -0.0158 | 0 | 1928 | FALSE | 4 | WM tract ICVF |
| IDP dMRI ProbtrackX ICVF atr l | BD | 0.8515 | [0.7897, 0.9181] | 2.868E-05 | 12 | -0.0147 | 6.912E-01 | 0.3477 | -0.0088 | 0 | 1952 | FALSE | 4 | WM tract ICVF |
| IDP dMRI TBSS ICVF Superior cerebellar peduncle R | BD | 1.1487 | [1.0634, 1.2409] | 4.326E-04 | 11 | 0.0087 | 8.143E-01 | 0.2979 | 0.0258 | 0 | 1914 | TRUE | 4 | WM tract ICVF |
| IDP T1 FAST ROIs L caudate | BD | 1.2114 | [1.0866, 1.3506] | 5.482E-04 | 9 | 0.062 | 1.136E-01 | 0.2696 | 0.0242 | 0 | 124 | FALSE | 4 | regional and tissue volume |
| BD | wg rh intensity-contrast insula | 0.0936 | [0.0309, 0.1564] | 3.459E-03 | 56 | -0.0142 | 7.280E-01 | 0.186 | 0.013 | 0 | 1436 | FALSE | 3 | cortical grey-white contrast |
| IDP dMRI TBSS ICVF Body of corpus callosum | BD | 0.9068 | [0.8592, 0.9572] | 3.888E-04 | 22 | -0.0356 | 2.812E-01 | 0.3707 | -0.0009 | -0.0123 | 1905 | FALSE | 3 | WM tract ICVF |
| IDP dMRI ProbtrackX ICVF slf l | BD | 0.8986 | [0.8443, 0.9563] | 7.591E-04 | 20 | -0.0561 | 1.093E-01 | 0.3737 | 0 | -0.004 | 1971 | FALSE | 3 | WM tract ICVF |
| IDP dMRI ProbtrackX ISOVF ifo r | BD | 0.8762 | [0.8158, 0.941] | 2.856E-04 | 16 | 0.0181 | 6.599E-01 | 0.2269 | -0.0135 | 0 | 2113 | FALSE | 3 | WM tract ISOVF |
| IDP dMRI TBSS ICVF Anterior corona radiata R | BD | 0.8846 | [0.8245, 0.9491] | 6.375E-04 | 15 | -0.0759 | 2.704E-02 | 0.3652 | -0.005 | 0 | 1924 | FALSE | 3 | WM tract ICVF |
| IDP dMRI TBSS ICVF Anterior limb of internal capsule L | BD | 0.8836 | [0.8193, 0.9528] | 1.306E-03 | 14 | 0.015 | 6.845E-01 | 0.344 | -0.0124 | 0 | 1919 | FALSE | 3 | WM tract ICVF |
| IDP dMRI TBSS ICVF Cerebral peduncle L | BD | 0.8731 | [0.8069, 0.9447] | 7.379E-04 | 13 | 0.1438 | 2.940E-04 | 0.301 | -0.0149 | 0 | 1917 | FALSE | 3 | WM tract ICVF |
| IDP dMRI ProbtrackX ICVF atr r | BD | 0.8614 | [0.7982, 0.9295] | 1.222E-04 | 12 | -0.0367 | 3.309E-01 | 0.3456 | -0.013 | 0 | 1953 | FALSE | 3 | WM tract ICVF |
| IDP dMRI ProbtrackX ICVF mcp | BD | 0.8581 | [0.7936, 0.9278] | 1.231E-04 | 11 | 0.0258 | 5.208E-01 | 0.2714 | -0.0175 | 0 | 1966 | FALSE | 3 | WM tract ICVF |
| IDP dMRI TBSS ICVF Anterior limb of internal capsule R | BD | 0.8581 | [0.7989, 0.9444] | 9.607E-04 | 11 | -0.012 | 7.386E-01 | 0.3524 | -0.0123 | 0 | 1918 | FALSE | 3 | WM tract ICVF |
| IDP dMRI TBSS MD Superior longitudinal fasciculus L | BD | 1.213 | [1.097, 1.3412] | 1.660E-04 | 9 | 0.0701 | 4.170E-02 | 0.3304 | 0.0206 | 0 | 1643 | FALSE | 3 | WM tract diffusivity |
| IDP dMRI ProbtrackX ICVF cst r | BD | 0.8578 | [0.787, 0.935] | 4.820E-04 | 9 | 0.012 | 7.409E-01 | 0.323 | -0.0114 | 0 | 1959 | FALSE | 3 | WM tract ICVF |
| IDP dMRI TBSS ICVF Cingulum hippocampus R | BD | 0.901 | [0.8472, 0.9582] | 9.025E-04 | 17 | -0.0028 | 9.435E-01 | 0.3054 | 0 | 0 | 1938 | FALSE | 2 | WM tract ICVF |
| IDP dMRI TBSS ICVF Posterior thalamic radiation L | BD | 0.9001 | [0.8445, 0.9595] | 1.246E-03 | 17 | -0.0973 | 1.087E-02 | 0.3058 | -0.002 | 0 | 1931 | FALSE | 2 | WM tract ICVF |
| IDP dMRI ProbtrackX L3 ifo r | BD | 1.1322 | [1.0462, 1.2253] | 2.064E-03 | 11 | 0.0693 | 5.886E-02 | 0.2951 | 0.0097 | 0 | 1888 | FALSE | 2 | WM tract diffusivity |
| IDP dMRI TBSS ICVF External capsule R | BD | 0.8906 | [0.8291, 0.9567] | 1.517E-03 | 11 | -0.0444 | 2.869E-01 | 0.3118 | -0.0066 | 0 | 1934 | FALSE | 2 | WM tract ICVF |
| IDP dMRI TBSS MD Anterior limb of internal capsule L | BD | 0.8383 | [0.7498, 0.9374] | 1.973E-03 | 8 | 0.0267 | 5.423E-01 | 0.2023 | -0.0294 | 0 | 1619 | FALSE | 2 | WM tract diffusivity |
